# Typically asymptomatic but with robust antibody formation: Children’s unique humoral immune response to SARS-CoV-2

**DOI:** 10.1101/2021.07.20.21260863

**Authors:** Hanna Renk, Alex Dulovic, Matthias Becker, Dorit Fabricius, Maria Zernickel, Daniel Junker, Alina Seidel, Rüdiger Groß, Alexander Hilger, Sebastian Bode, Linus Fritsch, Pauline Frieh, Anneke Haddad, Tessa Görne, Jonathan Remppis, Tina Ganzemueller, Andrea Dietz, Daniela Huzly, Hartmut Hengel, Klaus Kaier, Susanne Weber, Eva-Maria Jacobsen, Philipp D. Kaiser, Bjoern Traenkle, Ulrich Rothbauer, Maximilian Stich, Burkhard Tönshoff, Georg F. Hoffmann, Barbara Müller, Carolin Ludwig, Bernd Jahrsdörfer, Hubert Schrezenmeier, Andreas Peter, Sebastian Hörber, Thomas Iftner, Jan Münch, Thomas Stamminger, Hans-Jürgen Groß, Martin Wolkewitz, Corinna Engel, Marta Rizzi, Philipp Henneke, Axel R. Franz, Klaus-Michael Debatin, Nicole Schneiderhan-Marra, Ales Janda, Roland Elling

## Abstract

**Background:** Long-term persistence of antibodies against SARS-CoV-2, particularly the SARS-CoV-2 Spike Trimer, determines individual protection against infection and potentially viral spread. The quality of children’s natural humoral immune response following SARS-CoV-2 infection is yet incompletely understood but crucial to guide pediatric SARS-CoV-2 vaccination programs.

**Methods:** In this prospective observational multi-center cohort study, we followed 328 households, consisting of 548 children and 717 adults, with at least one member with a previous laboratory-confirmed SARS-CoV-2 infection. The serological response was assessed at 3-4 months and 11-12 months after infection using a bead-based multiplex immunoassay for 23 human coronavirus antigens including SARS-CoV-2 and its Variants of Concern (VOC) and endemic human coronaviruses (HCoVs), and additionally by three commercial SARS-CoV-2 antibody assays.

**Results:** Overall, 33.76% of SARS-CoV-2 exposed children and 57.88% adults were seropositive. Children were five times more likely to have seroconverted without symptoms compared to adults. Despite the frequently asymptomatic course of infection, children had higher specific antibody levels, and their antibodies persisted longer than in adults (96.22% versus 82.89% still seropositive 11-12 months post infection). Of note, symptomatic and asymptomatic infections induced similar humoral responses in all age groups. In symptomatic children, only dysgeusia was found as diagnostic indicator of COVID-19. SARS-CoV-2 infections occurred independent of HCoV serostatus. Antibody binding responses to VOCs were similar in children and adults, with reduced binding for the Beta variant in both groups.

**Conclusions:** The long-term humoral immune response to SARS-CoV-2 infection in children is robust and may provide long-term protection even after asymptomatic infection.

(Study ID at German Clinical Trials Register: 00021521)

## Introduction

To date, our knowledge of children’s immune response to infection with severe acute respiratory syndrome coronavirus type 2 (SARS-CoV-2) remains incomplete. In light of current debates on vaccination strategies and nonpharmaceutical preventative measures (e.g. school closures), a comprehensive understanding of protective immunity after natural infection in children is required. As with other viral infections, immune control of SARS-CoV-2 is achieved through a concerted interplay of humoral and cellular immunity (reviewed in ^1,2^). Neutralizing antibodies in children are of particular interest in this context, given their role in blocking virus entry into cells by inhibiting the interaction between the viral receptor binding domain (RBD) within the S-glycoprotein and the angiotensin-converting enzyme 2 (ACE2) receptor^3^.

Previous longitudinal studies of the humoral response have found that neutralizing antibodies peak within 3-5 weeks post-infection with a calculated half-life of up to 8 months, suggesting long-term protection in convalescent individuals^1,4–6^. However, most studies only included adults, and longitudinal studies on SARS-CoV-2 infections in children had limited sample size and duration of follow-up post-infection^7–17^. Furthermore, it remains unclear as to whether any form of cross-protection is offered by endemic human coronaviruses (HCoVs) that regularly circulate in the pediatric population, with some studies identifying cross-protection and others not.^18,19^

To provide an in-depth characterization of the humoral response in children, we initiated a multi-center longitudinal study, encompassing 328 households each with at least one SARS-CoV-2-infected member, which were followed for up to 12 months after the first infection in each household. This cohort is unique as the subjects exhibited mainly asymptomatic or mild disease with uninfected family members serving as environmental and age-matched controls. We performed an extensive serological evaluation of SARS-CoV-2 infection in all household members, comprising analyses of production of antibodies against various SARS-CoV-2 antigens, including Variants of Concern (VOCs), production of neutralizing antibodies and the role of endemic human coronaviruses (HCoVs).

## Materials and Methods

### Cohort

This study forms part of a non-interventional, prospective observational national multi-center cohort study, including 553 children and 726 adults from 328 households each with at least one individual with a SARS-CoV-2 reverse-transcriptase polymerase chain reaction (RT-PCR) proven infection and/or a symptomatic and later serologically proven infection. Participants were recruited during the first wave of the pandemic (May to August 2020) via local health authorities and an in-hospital database of households with at least one laboratory-confirmed SARS-CoV-2 infection. A full list of the inclusion and exclusion criteria are in the **Supplementary Appendix**.

### Study oversight

This part of the study was conducted by the University Children’s Hospitals in Freiburg, Tübingen and Ulm, Germany. Ethics approval was obtained from the respective Medical Faculties’ independent ethics committees (University of Freiburg: 256/20_201553; University of Tübingen: 293/2020BO2; University of Ulm: 152/20). Written informed consent was obtained from adult participants and from parents or legal guardians on behalf of their children at both sampling time points. Children’s preferences on whether or not to provide a blood sample were respected throughout. This study was registered at the German Clinical Trials Register (DRKS), study ID 00021521, conducted according to the Declaration of Helsinki, and designed, analyzed and reported according to the Strengthening the Reporting of Observational Studies in Epidemiology (STROBE) reporting guidelines.

### Sample collection

Samples were collected at two separate time points, an early time point (T1) at a median of 109 days (IQR 67-122 days) after earliest symptom onset in household and a late time point (T2) at 340 days (IQR 322-356 days) post-symptom onset (**Table 1, Figure S1 in Supplementary Appendix**).

**Table 1.** Demographics and key information for the study cohort.

| Timepoint<sup>a</sup> | T1<sup>a</sup> |  | T2<sup>a</sup> |  |
| --- | --- | --- | --- | --- |
| Number of participants by age group (n)<sup>a</sup> | Adult (717)<sup>a</sup> | Children (548)<sup>a</sup> | Adult (569)<sup>a</sup> | Children (402)<sup>a</sup> |
| Median Age (yr) (IQR)<sup>a</sup> | 44 (37-49)<sup>a</sup> | 10 (6-13)<sup>a</sup> | 45 (38-50)<sup>a</sup> | 10 (6-14)<sup>a</sup> |
| Number of females (%)<sup>a</sup> | 362 (50.49)<sup>a</sup> | 277 (50.55)<sup>a</sup> | 297 (52.20)<sup>a</sup> | 202 (50.25)<sup>a</sup> |
| Smoker (%)<sup>a</sup> | 75 (10.46)<sup>a</sup> | 2 (0.36)<sup>a</sup> | 62 (10.90)<sup>a</sup> | 2 (0.50)<sup>a</sup> |
| BMI (IQR)<sup>a</sup> | 25.37 (22.20-27.74)<sup>a</sup> | 17.41 (14.92-19.46)<sup>a</sup> | 24.69 (22.34-28.08)<sup>a</sup> | 16.96 (15.00-19.69)<sup>a</sup> |
| Seropositive (%)<sup>¶</sup> | 415 (57.88)<sup>¶</sup> | 185 (33.76)<sup>¶</sup> | 282 (49.56)<sup>¶</sup> | 151 (37.56)<sup>¶</sup> |
| - → Asymptomatic (%)<sup>¶</sup> | 36 (8.67)<sup>¶</sup> | 83 (44.86)<sup>¶</sup> | 31 (10.99)<sup>¶</sup> | 69 (45.70)<sup>¶</sup> |
| - → Symptomatic (%)<sup>a</sup> | 379 (91.33)<sup>a</sup> | 102 (55.14)<sup>a</sup> | 251 (89.01)<sup>a</sup> | 82 (54.30)<sup>a</sup> |
| Seroreverted at T2 (%)<sup>a</sup> | n/a<sup>a</sup> | n/a<sup>a</sup> | 71 (17.11)<sup>a</sup> | 7 (3.78)<sup>a</sup> |
| Symptoms at disease onset (of seropositive)<sup>¶</sup> |  |  |  |  |
| - → Fever (%)<sup>¶</sup> | 217 (52.29)<sup>¶</sup> | 66 (35.68)<sup>¶</sup> | 151 (53.55)<sup>¶</sup> | 49 (32.45)<sup>¶</sup> |
| - → Cough (%)<sup>¶</sup> | 221 (53.25)<sup>¶</sup> | 37 (20.00)<sup>¶</sup> | 154 (54.61)<sup>¶</sup> | 33 (21.85)<sup>¶</sup> |
| - → Dysgeusia (%)<sup>¶</sup> | 266 (64.10)<sup>¶</sup> | 28 (15.14)<sup>¶</sup> | 176 (62.41)<sup>¶</sup> | 24 (15.89)<sup>¶</sup> |
| - → Diarrhea (%)<sup>a</sup> | 75 (18.07)<sup>a</sup> | 18 (9.73)<sup>a</sup> | 55 (19.50)<sup>a</sup> | 16 (10.60)<sup>a</sup> |
| Median (IQR) days from positive PCR test result to timepoint<sup>a</sup> | 96 (63-120)<sup>a</sup> |  | 333 (319-353)<sup>a</sup> |  |
| Median (IQR) days from symptoms onset to timepoint (of seropositive)<sup>a</sup> | 109 (67-122)<sup>a</sup> |  | 340 (322-356)<sup>a</sup> |  |
| Hospitalised (of seropositive) (%)<sup>a</sup> | 15 (3.61)<sup>a</sup> | 0 (0.00)<sup>a</sup> | n/a<sup>a</sup> |  |
| Vaccinated (%)<sup>a</sup> | n/a<sup>a</sup> | n/a<sup>a</sup> | 24 (4.22)<sup>a</sup> | 1 (0.25)<sup>a</sup> |
| Number of households<sup>a</sup> | 328<sup>a</sup> |  | 279<sup>a</sup> |  |
| Median (IQR) number of household members<sup>a</sup> | 4 (3-4)<sup>a</sup> |  | 4 (3-4)<sup>a</sup> |  |

See methods for definition of how samples were defined as being seropositive, asymptomatic or symptomatic. Median time from positive PCR test to time point (n=368 at T1, n=310 at T2) and median time from symptoms onset to time point (n=349 at T1, n=243 at T2) are calculated using adult samples for which this data was available.

### Serological assays

Antibodies against SARS-CoV-2 in 2236 samples were detected using the following four assays: (1) EuroImmun-Anti-SARS-CoV-2 ELISA IgG and IgA (S1), (2) Siemens Healthineers SARS-CoV-2 IgG (RBD), (3) Roche Elecsys Ig (N) and (4) MULTICOV-AB, a previously published bead-based multiplex immunoassay that simultaneously analyses antibody binding to 23 antigens from SARS-CoV-2 (including VOCs)^20,21^. Seropositivity was defined as any three of the four SARS-CoV-2 assays being positive. The MULTICOV-AB assay also analyses antibody binding to endemic coronavirus antigens (i.e. HCoV-OC43, - NL63, -HKU1 and -229E)^20,21^. 385 samples were analyzed with the GenScript SARS-CoV-2 Surrogate Virus Neutralization Test (sVNT). See the **Supplementary Appendix** for details on the assay methodology.

### Data collection

Children and adults from eligible households were asked to provide information on demography and presence of symptoms (fever, cough, diarrhea or dysgeusia) in temporal relation to RT-PCR-proven or symptomatic and later serologically proven SARS-CoV-2 infection within the household. The four symptoms were chosen based on previous research and public health advice at the time. “Symptomatic infections” were defined by seropositivity and a history of at least one of these four symptoms in temporal relationship to exposure. Seropositivity in individuals who did not show any of these four symptoms was defined as “asymptomatic infection”. For most households, the earliest confirmed PCR-positive infection (“index case”) could be determined (see **Supplementary Appendix** for further information).

### Statistical Analysis

All data analysis was performed in RStudio (Version 1.2.5001), running R (version 3.6.1) with the additional packages “beeswarm”, “Rcolorbrewer”, “gplots” and “VennDiagram”. The type of statistical analysis performed and (where appropriate) the subset of the study population used is listed in the figure legends. Between-group differences of continuous endpoints were analyzed using Mann-Whitney-U tests while correlation was analyzed using the Spearman rank. All analyses were exploratory in nature and p-values may not be interpreted as confirmatory. A comprehensive material and methods section including detailed protocols can be found in the **Supplementary Appendix**.

## Results

548 children and 717 adults from 328 households were examined at T1 and 279 households including 402 children and 569 adults were followed to T2 (see Methods and Appendix for full details, **Table 1** for a description of the study population, **Figure S2** in the **Supplementary Appendix** for details on the age structure of the study population). Children were substantially less often seropositive (33.76% at T1, 37.56% at T2) than adults (57.88% at T1, 49.56% at T2) (**Table 1**). Seropositive participants were almost exclusively mildly or asymptomatically infected. In seropositive individuals, asymptomatic infections were five times more common in children (44.86% T1, 45.70% T2) than in adults (8.67% T1, 10.99% T2) (**Table 1**), with the proportion of asymptomatic infections decreasing with increasing age (**Figure S3**). Overall, hospitalization was rare (3.61% of adults, 0% of children, **Table 1**). The performance of the four serological assays for children and adults at T1 and T2 is shown in **Table S1 and Figure S4**.

The detailed humoral immune response against different SARS-CoV-2 antigens, assessed by MULTICOV-AB is shown in **Figure 1**. Children had significantly higher antibody titers against spike (p<0.001), RBD (p<0.001), S1 domain (p<0.001) and nucleocapsid (p=0.01) compared to adults at T1. This increased response was confirmed by the three commercial assays (**Figure S5**). In addition, we observed a large difference in seroreversion, with only 3.78% of children, but 17.11% of adults seroreverting between T1 and T2 (**Table 1**). Seroreversion was not associated with the response to particular antigens, although the largest and smallest decay in antibody concentrations were observed for antibodies against the S2 domain and nucleocapsid, respectively, regardless of age (**Figure S6**).

**Figure 1:**
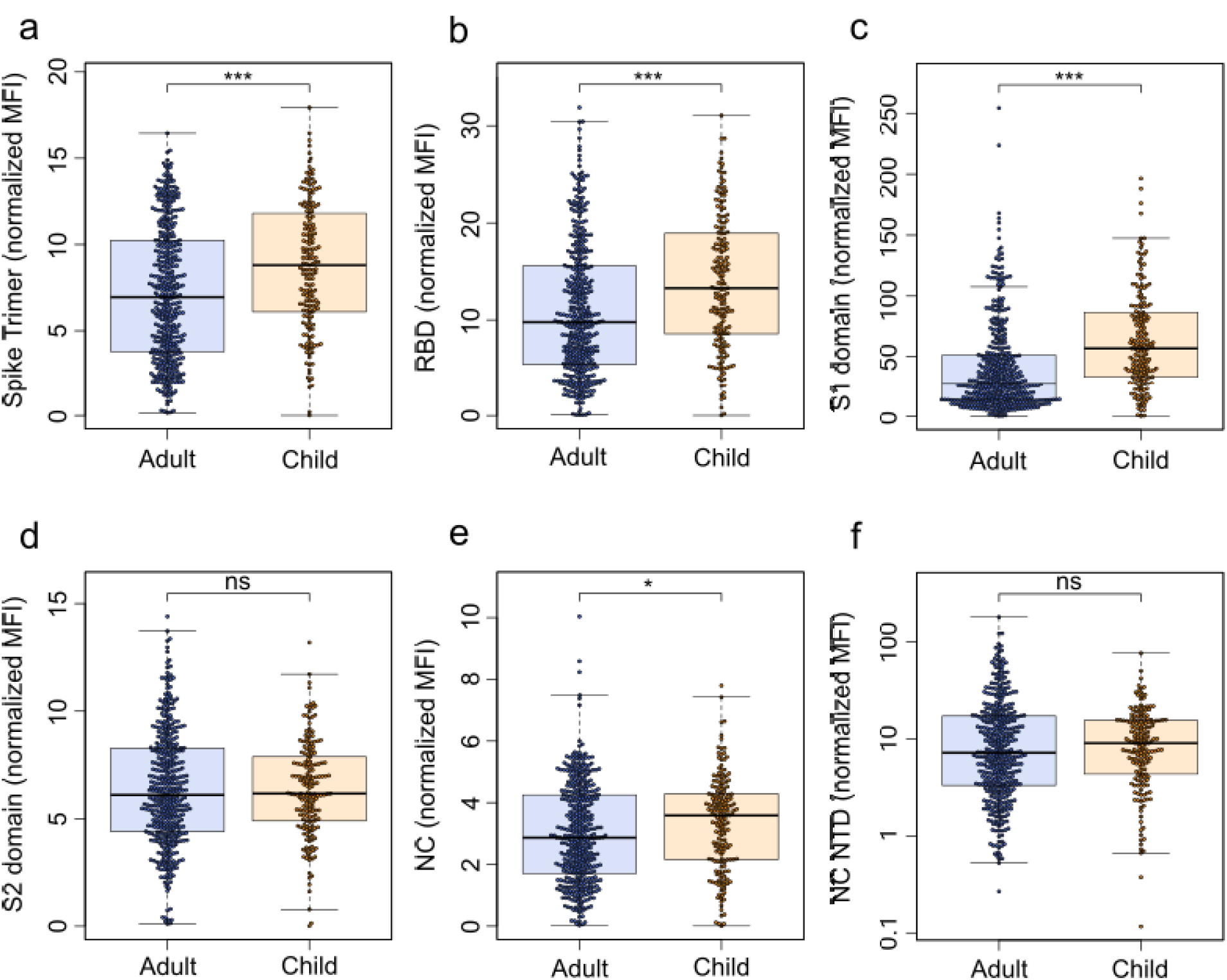
Children have a significantly higher humoral response to SARS-CoV-2 than adults. The humoral response generated following SARS-CoV-2 household exposure with seroconversion was examined using MULTICOV-AB. Children (n=181) produced significantly more antibodies against the Spike (**a**, p<0.001), RBD (**b**, p<0.001), S1 domain (**c**, p<0.001) and nucleocapsid (NC) (**e**, p=0.01) than adults (n=414). There was no significant difference for either the S2 domain (**d**, p=0.66) or the N-terminal domain of the nucleocapsid (NC NTD) (**f**, p=0.40). Only samples from T1 with a seropositive status (see Methods) are shown. Box and whisker plots with the box representing the median, 25th and 75th percentiles, while whiskers show the largest and smallest non-outlier values. Outliers were identified using upper/lower quartile ± 1.5 times IQR. Statistical significance was calculated using Mann-Whitney-U (two-sided) with significance defined as being *<0.01, ***<0.001

For both children and adults, there was no significant difference in antibody response between symptomatic and asymptomatic infections (**Figure 2a and b, Figure S7**). The frequency of reported symptoms differed between adults and children and the predictive value of each symptom varied between both groups (**Figure 2c and d**). While any of the symptoms fever, cough, diarrhea or dysgeusia proved to be a good indicator of infection in adults, dysgeusia was by far the best predictive symptom in children (87.50% of children with dysgeusia were seropositive; 95% CI 71.39%-95.15%, 30.52% of children without dysgeusia were seropositive for SARS-CoV-2, 95% CI 29.74%-31.30% **Figure 2d**). Conversely, cough was a poor predictor of SARS-CoV-2 infection in children (37.37% of children with a cough were seropositive; 95% CI 29.25%-46.28%, 33.04% of children without a cough were seropositive; 95% CI 30.99%-35.15%, **Figure 2d**). Further examination of predictive symptoms among children showed that in contrast to dysgeusia, cough only gained predictive value in children above the age of 12 and the predictive value of fever increased with age (**Table S2**). There was no difference in the humoral response associated with the presence of particular symptoms in either adults or children (**Figure S8**).

**Figure 2:**
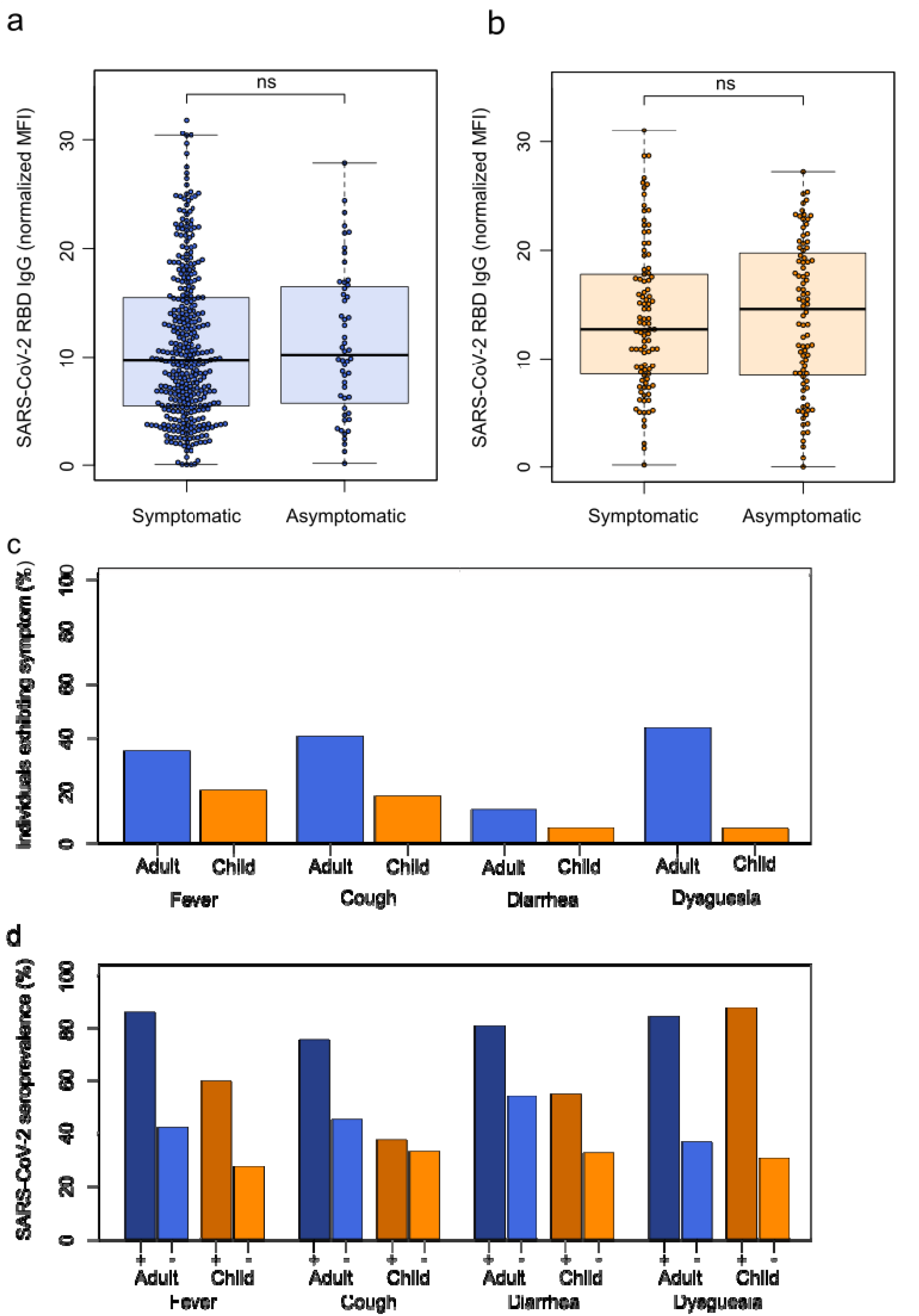
SARS-CoV-2 infections in children are more often asymptomatic than in adults, although dysgeusia is a good indicator of SARS-CoV-2 infection in both adults and children. Box and whisker plots showing that there is no difference in antibody response between asymptomatic and symptomatic SARS-CoV-2 infections in adults (**a**, p=0.684, n=415) or children (**b**, p=0.712, n=185). Boxes represent the median, 25th and 75th percentiles, while whiskers show the largest and smallest non-outlier values. Outliers were identified using upper/lower quartile ± 1.5 times IQR. Statistical significance was calculated using Mann-Whitney-U (two-sided). The four symptoms reported in this study were then examined for their frequency within the study population (**c**), with all symptoms more commonly reported in seropositive adults than seropositive children. Each symptom was then examined for its predictive ability to indicate SARS-CoV-2 infection (**d**), with dysgeusia a strong predictor in both adults (84.18%) and children (87.50%). All other symptoms were poor predictors in children (fever 59.46%, cough 37.37%, diarrhea 54.55%) compared to adults (fever 85.77%, cough 75.03%, diarrhea 80.65%). Only samples from T1 were analyzed for this figure (n=717 adults, 548 children). “+” indicates presence of the symptom “-“ indicates absence of the symptom.

To further explore differences in the antibodies produced by children and adults, we analyzed their neutralization potential in a surrogate assay (sVNT) as well as their binding towards VOCs. The neutralizing potential of children’s sera exceeded that of adults’ at T1 (p<0.001) and T2 (p=0.02) (**Figure 3a**). However, this could be attributed to antibody titers, as neutralization in children correlated with the S1-directed antibody response (Spearman’s rank 0.86, **Figure 3b**). There was no difference in antibody binding responses to the RBD of Alpha and Beta VOCs between adults and children, with an identical binding for the Alpha variant compared to wild-type (Spearman’s rank 0.95, **Figure 3c**) and a reduction in binding for the Beta variant (Spearman’s rank 0.69, **Figure 3d**).

**Figure 3.**
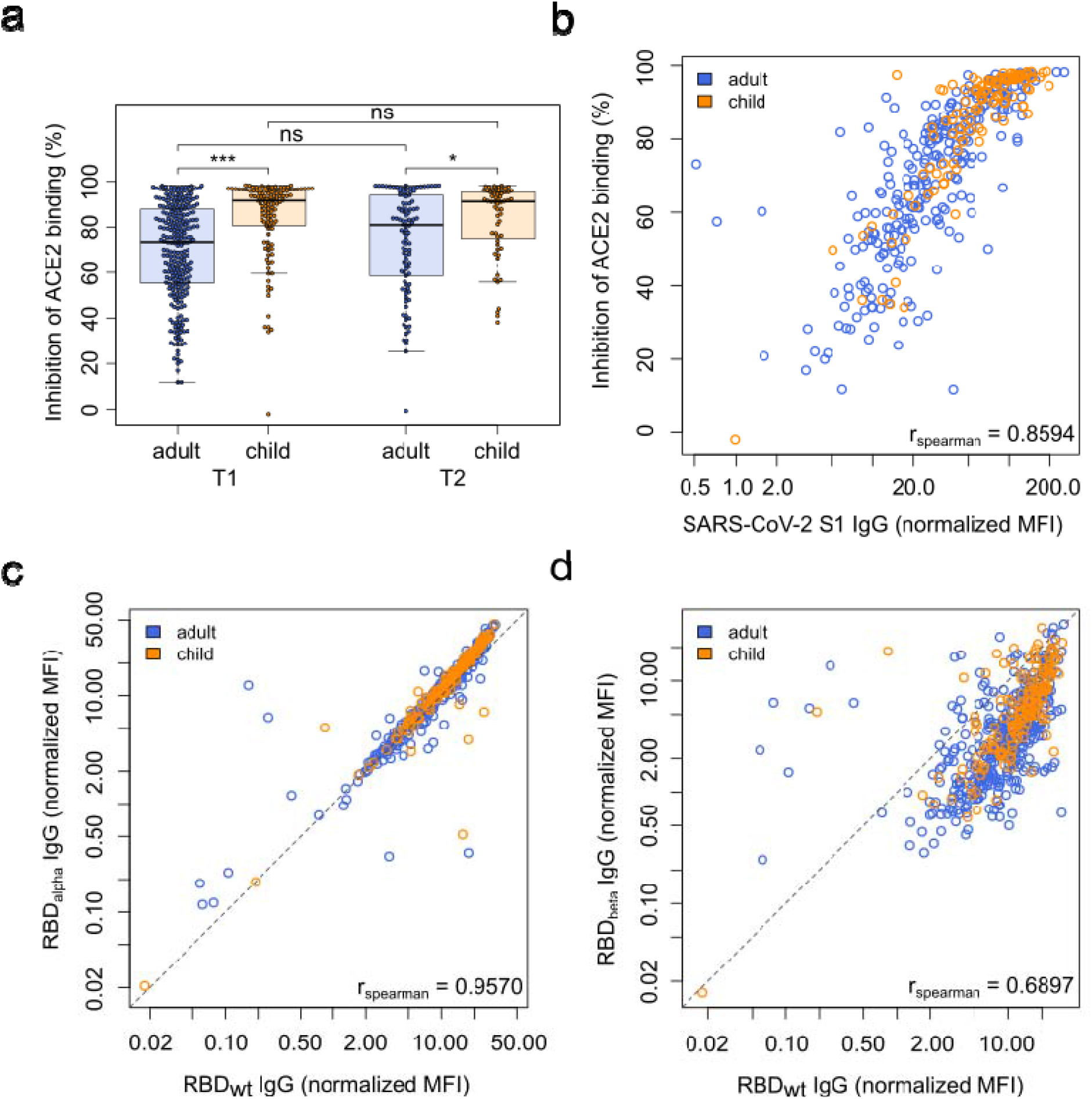
Children and adults produce antibodies with equal neutralizing potential and their antibodies offer the same protection against Variants of Concern. (**a**) Box and whisker plot showing that antibodies produced by children (n=118) have a significantly higher inhibition of ACE2 binding than those produced by adults (n=267, p<0.001) at T1 and T2 (p=0.02, child n=59, adult n=106) as determined by the sVNT assay. Boxes represent the median, 25th and 75th percentiles, while whiskers show the largest and smallest non-outlier values. Outliers were identified using upper/lower quartile ± 1.5 times IQR. Statistical significance was calculated using Mann-Whitney-U (two-sided) with *** indicating a p-value <0.001. To determine whether this was due to the higher titers in children, SARS-CoV-2 S1 humoral response was determined using MULTICOV-AB for T1 and plotted against the results of the sVNT assay, with Spearman’s rank used to determine the correlation (**b**), confirming that the increase in neutralization is due to higher titers. Protection against the Alpha (**c**) and Beta (**d**) VOCs was determined by MULTICOV-AB and plotted as a linear regression against the antibody binding response to the wild-type (wt) RBD, with Spearman’s rank used to determine the correlation. There was no difference in antibody response between children (n=166, T1 samples only) and adults (n=381, T1 samples only) for either variant.

Seroprevalence against endemic coronaviruses rose sharply with age in early childhood, and was stable in older children, adolescents and adults independent of age (**Figure 4a, Figure S9**). In contrast to SARS-CoV-2 seroreversion, HCoV antibody titers decreased faster in younger children than in adults (**Figure S10**). There were HCoV naïve samples in this cohort and some individuals showed a substantial increase in HCoV antibody response indicating exposure towards endemic HCoVs between the two time points (**Figure 4b in red, Figure S11**). Amongst SARS-CoV-2 exposed individuals in households with a defined index case (index cases excluded from the analysis, see Methods), there was no difference in HCoV antibody titers between SARS-CoV-2 seropositive and seronegative children or adults (p=0.21, **Figure 4c, Figure S12**). In addition, we assessed whether SARS-CoV-2 infection boosted HCoV antibody responses, however there was no evidence for an association between HCoV antibody responses and SARS-CoV-2 antibody responses in exposed children or adults (Spearman’s rank 0.03, **Figure 4d**).

**Figure 4:**
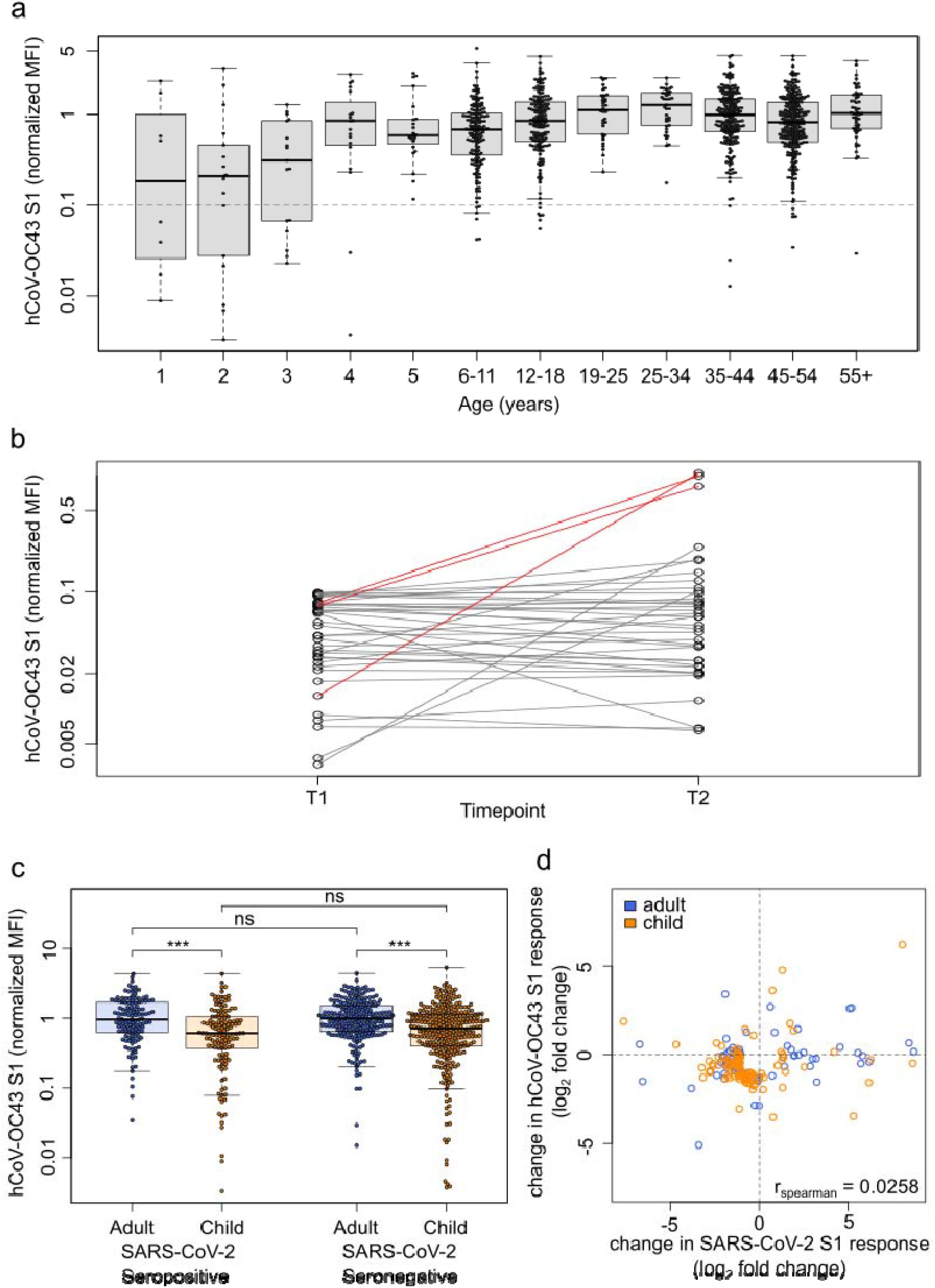
HCoVs offer no protection against SARS-CoV-2, nor do they show a boost-back antibody response following SARS-CoV-2 infection. Samples from households with a known index case were examined with MULTICOV-AB to determine whether the antibody response to endemic coronaviruses (HCoV) provides any protection against infection with SARS-CoV-2. Initial screening of the population showed that seroprevalence increases with age, although several samples were within the blank range of the HCoV assays, indicating the presence of naïve samples (**a**). Naïve samples were defined as those having less than one-tenth the mean antibody response (indicated by dotted line), with the majority of these samples occurring in children under the age of 5. (**b**) Line graph showing the longitudinal response of these naïve samples from T1 to T2, with new infections in HCoV-OC43 shown in red. (**c**) Box and whisker plot showing there is no significant difference in HCoV-OC43 antibody response between SARS-CoV-2 seropositive and seronegative individuals, among either adults (n=440, p=0.974) or children (n=436, p=0.214). Boxes represent the median, 25th and 75th percentiles, while whiskers show the largest and smallest non-outlier values. Outliers were identified using upper/lower quartile ± 1.5 times IQR. Statistical significance was calculated by Mann-Whitney-U (two-sided) with *** indicating a p-value <0.001 and ns indicating a p-value >0.01. (**d**) When comparing paired samples longitudinally within the SARS-CoV-2 seropositive subgroup, there was no increase in HCoV-OC43 S1 response in either adults (n=76) or children (n=103) following SARS-CoV-2 infection. Change in response is presented as log2-fold change from T1 to T2 and only samples with either log2-fold change greater than 1 or smaller than -1 are shown. Spearman’s rank was used to calculate any correlation between the change in response for HCoV-OC43 and SARS-CoV-2. The same figures for the endemic coronaviruses HCoV-NL63, HCoV-HKU1 and HCoV-229E can be found as Supplementary figures 10 – 12.

## Discussion

To our knowledge, this is the largest prospective multi-center study comprehensively comparing the adult and pediatric longitudinal humoral immune response following SARS-CoV-2 household exposure. As the humoral immunity against SARS-CoV-2 is now increasingly accepted as the central correlate of protection^22–24^, improving our incomplete understanding in children^25,26^ is of considerable value for public health and vaccination strategies. Importantly, our outpatient cohort has high epidemiological relevance, as a mild course is the most frequent outcome of SARS-CoV-2 infection overall^27^. Our findings identify several unique features of the pediatric serological immune response against SARS-CoV-2.

Children had a lower seroprevalence after household exposure and seropositivity followed asymptomatic infection more frequently than in adults. This is in agreement with our previous report of a different cohort consisting of parent-child pairs^28^. In light of potential pediatric vaccination campaigns, children’s humoral response to SARS-CoV-2 is markedly increased in both quantity and longevity, with children seroreverting significantly slower than adults. Children generated a higher titers of SARS-CoV-2 antibodies than their parents after being exposed to the same viral strain, and antibody titers are negatively correlated with age. Of particular interest are the increase in antibodies produced against the S1 domain and RBD, both of which are associated with higher neutralization capacity, indicating that children produce a high quality humoral response against SARS-CoV-2 ^3,22,29^. The quality of the pediatric humoral response is further illustrated by the similar binding capacity against the SARS-CoV-2 Alpha and Beta VOCs compared to adults. Children also had significantly higher neutralizing antibody titers than adults, indicating increased protection. This increase in neutralization was directly correlated with higher antibody titers in the other assays, and therefore may not be due to substantial qualitative changes of the pediatric antibody profiles. These findings are in line with one preprinted study^30^ but in contrast to two previous studies, which found that children generated a lower humoral response to SARS-CoV-2 than adults, with a corresponding reduction in neutralization activity^11,13^. However, compared to our cohort, all three studies were substantially smaller in sample size and the latter two investigated a different disease spectrum comprising mostly hospitalized children or those diagnosed with hyperinflammatory MIS-C syndrome, and sampled blood at earlier time points after presumed infection.

It is striking that antibody levels in seropositive individuals were independent of fever, cough or diarrhea, as clinical proxies for systemic or localized inflammation of the respiratory or gastrointestinal tract, respectively. Previous studies have reported a clear correlation between disease severity and neutralizing antibody titers in adults^6,31^. At the other end of the disease spectrum with mildly affected younger adults and children, this association was not detectable irrespective of age. This diverges from the classical infection immunology dogma that systemic pathogen-host interaction is required for the generation of robust immune memory. While titers themselves did not differ between asymptomatic and symptomatic infections, we found substantial differences in titers between adults and children. Presence of any symptom was predictive of seropositivity in adults, whereas children showed substantial differences in both the prevalence of symptoms in seropositive individuals and the predictive values of symptoms with respect to SARS-CoV-2 seropositivity. Since cough was a relatively common symptom in children irrespective of seroconversion, it was not useful in predicting SARS-CoV-2 infection. In contrast, dysgeusia, an infrequent symptom among children, was highly accurate in predicting infection. These findings suggest that symptom criteria used for subsequent PCR testing need to be different for children and adults.

Similarly to other authors^32,33^, we identified that exposure to HCoVs, as measured by seropositivity typically happens within the first five years of life. The relatively small decline of HCoV antibody levels during the study period, especially in the adult population, in comparison to the decline in SARS-CoV-2 antibody levels after a single infection suggests that long-term serological immunity against HCoVs may be driven by recurrent exposure. We observed HCoV infections in previously naïve individuals, indicating that endemic HCoVs still circulated between T1 and T2 despite SARS-CoV-2 related distancing and hygiene measures. Although cross-reactivity and/or cross-protection between SARS-CoV-2 and HCoVs have been hypothesized, our analyses did not find evidence for such effects. For both Alpha- and Beta-coronaviruses, HCoV antibody responses were not associated with a lower likelihood of seroconversion following SARS-CoV-2 exposure. Along with frequent HCoV seronegativity in younger childhood, this strongly argues that the lower incidence of SARS-CoV-2 infection in children is not due to HCoV cross-protection. Moreover, there was no evidence for boosting of HCoV titers following SARS-CoV-2 infection. In contrast to other studies which did identify an effect for endemic HCoV infection, our cohort is composed of intensely exposed individuals from within the same households, which is a substantial strength compared to previous studies that have used pre-pandemic sera or indirect control groups^18,30,24^.

Limitations of our study include the potential recall-bias inherent to retrospective self- or parent-reporting of symptoms via questionnaires and physician-interviews. Additionally, PCR tests for SARS-CoV-2 during the first wave in Germany were mostly limited to the household index case, meaning it is possible that infected individuals were not identified as such, despite the multi-assay serological approach. However, the in-depth characterization of the humoral response provides valuable data for clinicians, public health officials and the public, at a time when children are increasingly viewed as a potential viral reservoir due to exclusion of pediatric populations from current vaccination strategies. Similarly, while PCR testing was not available for all individuals, the strength of this cohort comes from the comparatively large number of children, inclusion of children and adults from the same household, the inclusion of seronegative household members as well-matched controls, and the prospective longitudinal analysis of the humoral response in children for up to one year post-infection. The seropositive cohort also comprises almost exclusively individuals with mild or asymptomatic infections and so provides real-world data representative for the majority of SARS-CoV-2 infections in the community.

In summary, although children mostly show mild or even asymptomatic clinical courses following SARS-CoV-2 infection, they mount a strong and enduring humoral immune response. This strongly argues for sustained protection after infection, and might inform the design of vaccination strategies for SARS-CoV-2 convalescent children.

## Data Availability

Data from this study are available upon request.

## Author Contributions

RE, HR, AJ, DF, PH, ARF and KMD conceived the study. HR, ADu, RE, AJ, PH, ARF, KMD and NSM designed the experiments. RE, HR, AJ, PH, ARF, KMD and NSM procured funding. ADu, MB, DJ, AS, RG, JM, AHi, CL, TG, ADi, DH, HH, AP, TI, TS and H-JG performed experiments. RE, HR, AJ, DF, MZ SB, LF, PF, AHa, JR, E-MJ, CE, MW, TG and MR collected samples or organized their collection. BJ, HS, MS, BT, GFH and BM supported the sample collection and provided key resources. PK, BT and UR produced the RBD mutants. HR, AJ, ADu, MB, DF, AHi, ADi, KK, SW, E-MJ, AP, TI, TS, H-JG, MW, CE, KMD and MR curated the data. MB and ADu performed the data analysis. ADu and MB generated the figures. ADu, HR, AJ and RE wrote the first draft of the manuscript. All authors approved the final version of the manuscript.

## Acknowledgements

We thank Carmen Blum, Sevil Essig, Ulrike Formentini, Jens Gruber, Andrea Hänsler, Simone Hock, Ann Kathrin Horlacher, Jennifer Juengling, Gudrun Kirsch, Ingrid Knape, Helgard Knauss, Sonja Landthaler Alexandra Niedermeyer, Bianca Rippberger, Andrea Schuster, Boram Song, Ulrike Tengler, Mareike Walenta and Linda Wolf for assistance with sample processing and patient material storage. We are grateful for the FREEZE and HILDA biobank Freiburg for sample processing, in particular Ali-Riza Kaya; Marco Teller and Dirk Lebrecht. We thank Sandra Steinmann, Yvonne Müller, Vanessa Missel, Andrea Evers-Bischoff, Andrea Bevot and the CPCS at the University Hospital Tübingen for organizational support in conducting the study. We thank Steffen Keul for assistance with data processing. This work was financially supported by the State Ministry of Baden-Württemberg for Economic Affairs, Labour and Housing Construction (grant numbers FKZ-3-4332.62-NMI-67 and FKZ-3-4332.62-NMI-68) to NSM and the Ministry of Science, Research and the Arts Baden-Württemberg within the framework of the special funding line for COVID-19 research to the Freiburg, Tübingen, Ulm and Heidelberg centers. The funders had no role in study design, data collection, data analysis or the decision to publish.

