## Supplement for "Typically asymptomatic but with robust antibody formation: Children’s unique humoral immune response to SARS-CoV-2"

### Supplementary Appendix

**Supplementary Contents**

#### List of investigators

Hanna Renk MD[1*], Alex Dulovic Dr.rer.nat[2*], Matthias Becker M.Sc[2], Dorit Fabricius MD[3], Maria Zernickel[3], Daniel Junker M.Sc[2], Alina Seidel M.Sc[4], Rüdiger Groß M.Sc[4], Alexander Hilger M.Sc[5], Sebastian Bode MD[3], Linus Fritsch[5], Pauline Frieh[5], Anneke Haddad DPhil[5], Tessa Görne[5], Jonathan Remppis MD[1], Tina Ganzemueller MD[7], Andrea Dietz Dr.biol.hum[6], Daniela Huzly MD[8], Hartmut Hengel MD[8], Klaus Kaier PhD[9], Susanne Weber PhD[9], Eva-Maria Jacobsen PhD[3], Philipp D. Kaiser Dr.rer.nat[2], Bjoern Traenkle Dr.rer.nat[2], Ulrich Rothbauer Dr.rer.nat[2], Maximilian Stich MD[10], Burkhard Tönshoff[10], Georg F. Hoffmann[10], Barbara Müller PhD[10,11], Carolin Ludwig[12,13,14], Bernd Jahrsdörfer MD[12,13,14], Hubert Schrezenmeier MD[12,13,14], Andreas Peter MD[15], Sebastian Hörber MD[15], Thomas Iftner PhD[7], Jan Münch PhD[4], Thomas Stamminger MD[6], Hans-Jürgen Groß MD[16], Martin Wolkewitz PhD[9], Corinna Engel Dr.biol.hum[1,17], Marta Rizzi MD[18], Philipp Henneke MD[5,19], Axel R. Franz MD[1,17], Klaus-Michael Debatin MD[3], Nicole Schneiderhan-Marra Dr.rer.nat[2], Ales Janda MD[3,#] and Roland Elling MD[5,19,#,†]

1 – University Children's Hospital Tübingen, Tübingen, Germany

2 – NMI Natural and Medical Sciences Institute at the University of Tübingen, Reutlingen, Germany

3 – Department of Pediatrics and Adolescent Medicine, Ulm University Medical Center, Ulm University, Ulm, Germany

4 – Institute of Molecular Virology, Ulm University, Ulm, Germany

5 – Center for Pediatrics and Adolescent Medicine, Medical Center Freiburg, Germany and Faculty of Medicine, University of Freiburg, Freiburg, Germany

6 – Institute of Virology, Ulm University Medical Center, Ulm, Germany

7 – Institute for Medical Virology and Epidemiology of Viral Diseases, University Hospital Tübingen, Tübingen, Germany

8 – Institute of Virology, University Medical Center Freiburg, Germany and Faculty of Medicine, University of Freiburg, Freiburg, Germany

9 – Institute of Medical Biometry and Statistics, Faculty of Medicine and Medical Center, University of Freiburg, Freiburg, Germany

10 – Department of Pediatrics I, University Children's Hospital Heidelberg, Heidelberg, Germany

11 - Department of Infectious Diseases, Virology, Heidelberg University, Heidelberg, Germany

12 – Department of Transfusion Medicine, Ulm University, Ulm, Germany

13 – Institute for Clinical Transfusion Medicine and Immunogenetics

14– German Red Cross Blood Transfusion Service, Baden-Württemberg, Germany

15 – Institute for Clinical Chemistry and Pathobiochemistry, University Hospital Tübingen, Tübingen, Germany

16 – Institute of Clinical Chemistry, Ulm University, Ulm, Germany

17 – Center for Pediatric Clinical Studies, University Hospital Tübingen, Tübingen, Germany

18 - Department of Rheumatology and Clinical Immunology, Freiburg University Medical Center, Faculty of Medicine, University of Freiburg, Freiburg, Germany

19 – Institute for Immunodeficiency, University Medical Center Freiburg, Freiburg, Germany

*indicates shared first authorship

#indicates shared last authorship

[†]indicates corresponding author

#### Supplementary Methods

Study Design

We did a secondary analysis of a non-interventional, prospective observational national multi-center cohort study, including 548 children and 717 adults within 328 households with at least one SARS-CoV-2 RT-PCR-confirmed or one seropositive, symptomatic individual. Due to restrictions in obtaining a SARS-CoV-2 RT-PCR test during the first wave (children and asymptomatic contacts of an index case were not tested routinely), serological assays were the only means to identify previous infection. This study was initiated by the four University Children's Hospitals of Freiburg, Heidelberg, Tübingen and Ulm and approved by the independent ethics committees of each center. Sera and data for this substudy were collected at the study sites in Freiburg, Tübingen and Ulm. Participants were asked to fill a questionnaire at time point 1 (May– August 2020) and follow-up time point 2 (February-March 2021).

Study Participants and Eligibility Criteria

Families were identified during the first wave of the pandemic between May and August 2020 in the region of Baden-Württemberg, Germany.

Inclusion and exclusion criteria were as follows:

Subjects were eligible for enrolment if they met the following inclusion criteria:

(i) Children (male or female) aged 1 to 18 years,

(ii) parents and other adults (male or female) living in the same household with the investigated children (without age limit),

(iii) residency in the state of Baden-Württemberg,

(iv) written consent to the study.

Key exclusion criteria were

(i) severe congenital diseases (e.g. infantile cerebral palsy, severe congenital malformations)

(ii) congenital or acquired immunodeficiencies,

(iii) insufficient comprehension of German.

Data Collection

Children and adults within eligible households completed a questionnaire containing demographic information (date of birth, gender, height, weight, smoking), the presence of symptoms (fever, cough, dysgeusia or diarrhea) in plausible temporal association (max. 2 weeks prior or later) with the onset of the SARS-CoV-2 infection within the household or around the time of a positive SARS-CoV-2 RT-PCR, and symptom duration. They additionally provided serum samples for immunological analysis at time point 1 and at follow-up time point 2 after the SARS-CoV-2 infection within the household. Data on vaccination and potential re-infection within the household were collected at time point 2. We investigated all invited households with at least one child to avoid selection bias. Questionnaires were checked for missing or inadequate data and inconsistencies; where possible, these points were clarified retrospectively with the families. To predetermine the sample size, we used a one-factor variance analysis design. Assuming 1.5 children per household participating in the study and 3 different age ranges, we aimed at a sample size of approximately 200 households to reveal small effect sizes of about 0.1 at a significance level of 5% and a test strength of 80%.

Data variables

Samples were defined as being “symptomatic” on the basis of having at least one of four (fever, cough, diarrhea and dysguesia) symptoms, in addition to a positive serology result. “Asymptomatic” samples were defined as those without any of the above symptoms and a positive serology result. For some households (n=272) an “index case” was defined. This is the first household member to test positive for SARS-CoV-2 by RT-PCR. No index was defined for households where additional household members tested positive by RT-PCR within 48 hours of the first positive test or where infection was identified by the combination of positive serology and symptoms only. If a household had a defined index case, then all other household members were considered “exposed”. “Time post symptom onset” within a household was calculated as the number of days between the first date of symptom onset in any seropositive individual within a household and the sampling date at T1 and T2 respectively.

Blood Sample Collection

Blood samples were collected by venipuncture from all consenting adults and children within the study. Serum was separated on the same day by centrifugation, aliquoted and frozen at -80°C until used.

EuroImmun Anti-SARS-CoV-2 ELISA

The EuroImmun Anti-SARS-CoV-2 ELISA (IgA and IgG) was performed as the manufacturer’s instructions to detect IgG or IgA antibodies against the S1 domain of the SARS-CoV-2 spike protein. All 2236 samples used in the final analysis were measured with this assay. All samples were processed with the specified controls and calibrators. Serological analysis was performed blinded for all clinical covariables.

Siemens Healthineers SARS-CoV-2 IgG (sCOVG)

The Siemens sCOVG assay was performed as per the manufacturer’s instructions on an Advia Centaur XPT platform to detect IgG antibodies against the receptor-binding-domain (RBD) of the SARS-CoV-2 spike protein. All 2236 samples used in the final analysis were measured with this assay. All samples were processed with the specified controls and calibrators. Serological analysis was performed blinded for all clinical covariables.

Roche Elecsys Electrochemiluminescence immunoassay (ECLIA)

The Roche Elecsys ECLIA was performed as per the manufacturer's instruction on a Cobas e411 or e811 platform to detect IgG, IgA and IgM antibodies against the nucleocapsid of SARS-CoV-2. All 2236 samples used in the final analysis were measured with this assay. All samples were processed with the specified controls and calibrators. Serological analysis was performed blinded for all clinical covariables.

MULTICOV-AB™

All 2236 samples used in the final analysis were analyzed using MULTICOV-AB™[1], a bead-based multiplex immunoassay that simultaneously analyzes 23 antigens from SARS-CoV-2 (including RBDs from variants of concern and endemic human coronaviruses)[1]. A full list of antigens used in this study can be found in **Table S3**. Samples were measured in 384-well plates, with all pipetting steps performed using a Beckmann Coulter i7 pipetting robot. Antigens were coupled by EDC/s-NHS or Anteo coupling to spectrally distinct populations of MagPlex beads (Luminex Technology). Samples were diluted in assay buffer (1:4 Low Cross Buffer (Candor Bioscience GmBH) in CBS (1x PBS + 1% BSA) + 0.05% Tween20) and added to bead mix to a final dilution factor of 1:400, before being incubated for 2 hours at 21°C on a thermomixer (1500rpm). Unbound antibodies were then removed by washing with Wash buffer (1x PBS, 0.05% Tween20). Bound antibodies were detected using RPE-conjugated human IgG (3 µg/mL) and IgA (5 µg/mL) (both Biozol) by incubation for 45 mins at 21°C, 1800 rpm on a thermomixer. Following a further washing step, beads were resuspended in 80µL of washing buffer and shaken briefly for 3 mins at 1500 rpm. Plates were then measured using a FLEXMAP-3D (Luminex Technology) instrument with the following settings: 60 µL, 80 s timeout, 35 events, Gate 7500-15000 and Reporter Gain: Standard PMT. As quality control, three QC samples with known Median Fluorescence Intensity (MFI) values were included on each plate.[2,3] Eight wells for each QC sample plus 8 blank wells (negative control) were included on each 384-well plate. Additionally, control beads coupled with human IgG, goat-anti-human IgG, human IgA and goat-anti-human IgA were included in each well to act as controls for both sample addition and signal system addition. To pass QC, each sample had to meet the minimum threshold for number of beads per ID (35), have a sample and signal system control bead value within normal range and pass plate-by-plate QC sample controls. Any plate or sample that failed QC was re-measured (83/2390). Normalization values for each antigen were generated by dividing the raw median fluorescence intensity (MFI) value by the mean plate-by-plate MFI of QC2 (IgG) or QC3 (IgA). For SARS-CoV-2, normalization values >1 for the trimeric spike and wild-type RBD indicate positivity. To reduce analytical variations, all samples were analyzed in the same run. Serological analysis was performed blinded for all clinical covariables. Technical questions regarding the MULTICOV-AB™assay should be directed to.

Surrogate SARS-CoV-2 Neutralization Test

A subset of 385 samples were analyzed for neutralization with the surrogate SARS-CoV-2 neutralization test (GenScript) as per the manufacturer's instructions and as published previously[4]. Briefly, samples and controls were incubated with an HRP-conjugated RBD fragment. Following this, the mixture was added to wells of a capture plate coated with human ACE2 protein. The plate was then washed three times to remove any complexes or non-bound antibodies. TMB was added and then stopped with the addition of a stop reagent. The plate was then read by a microtiter plate reader at 450 nm. The absorbance of the sample is inversely correlated with the amount of SARS-CoV-2 neutralizing antibodies. Positive and negative controls served as internal assay quality controls. The test was considered valid only if the OD450 for each control fell within the respective range (OD450negative control > 1.0, OD450positive control < 0.3). For final interpretation, inhibition rates were calculated as follows: Inhibition score (%) = (1 - (OD valuesample/OD valuenegative control) x 100%). Scores < 30% were considered negative, scores ≥ 30% were considered positive.

Data Analysis

Initial data collection was done using Microsoft Excel and Access. Formal data analysis was performed on RStudio (Version 1.2.5001, running R 3.6.1) with the following additional packages: “RColorBrewer”, “beeswarm”, “gplots”, “VennDiagram”, all of which were used solely for data depiction and not statistical analysis. Figures were generated in RStudio and then edited for clarity in Inkscape (Inkscape 0.92.4). Only samples for which full data for MULTICOV-AB was available were included in the analysis. Furthermore, only samples from time point 1 were used for all non-longitudinal analyses. For longitudinal analyses, only those participants for whom both T1 and T2 samples were available were included and all participants who were vaccinated prior to T2 were excluded. For analysis of potential cross-protection though endemic coronaviruses, only households with a known index case were used and the index case itself was excluded. Statistical analyses performed are described in the figure legends. For comparison of signal distribution between sample groups, Mann-Whitney-U tests were performed using the “wilcox.test” function from R’s “stats” library. For correlation analysis, Spearman’s rank was calculated using the “cor” function from R’s “stats” library. p-values <0.01 were considered to be significant.

#### Supplementary Figures

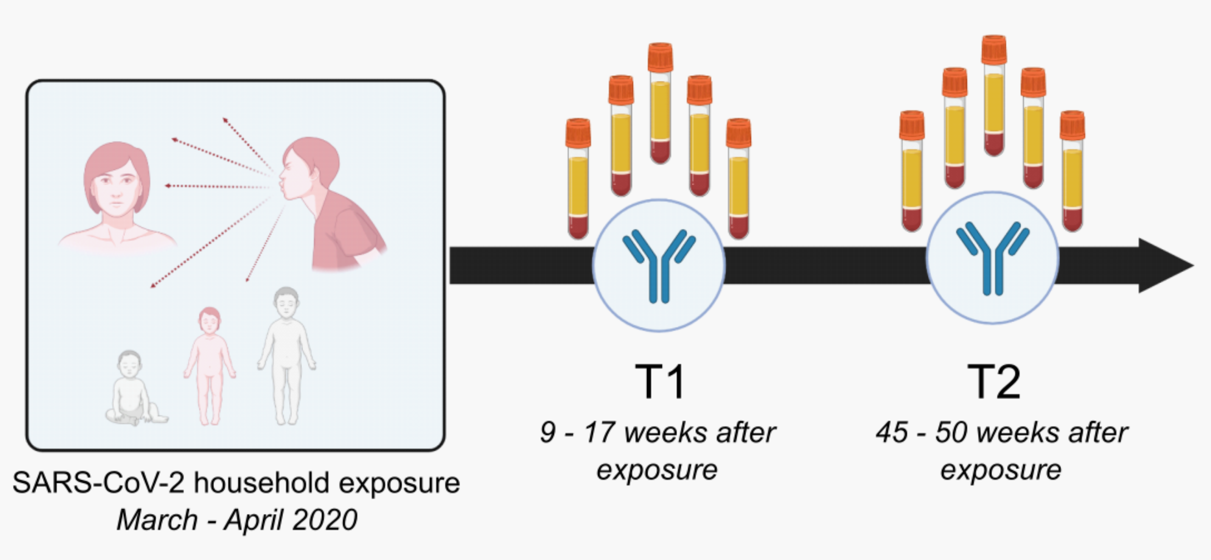

Figure S1: **Overview of the time points within the study population.** Illustration of study design, from exposure to study participation time points. Times shown are the IQR for each time point.

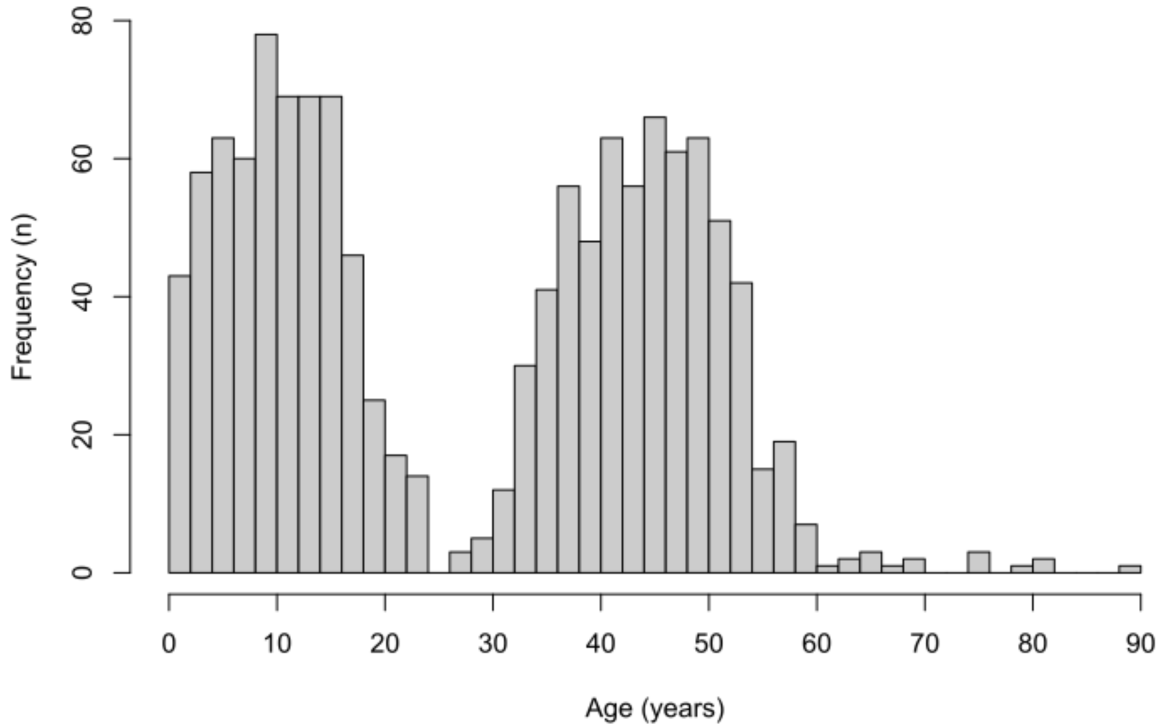

Figure S2 – **Study population age distribution**. Histogram showing age distribution within the study population at T1 (n=1265).

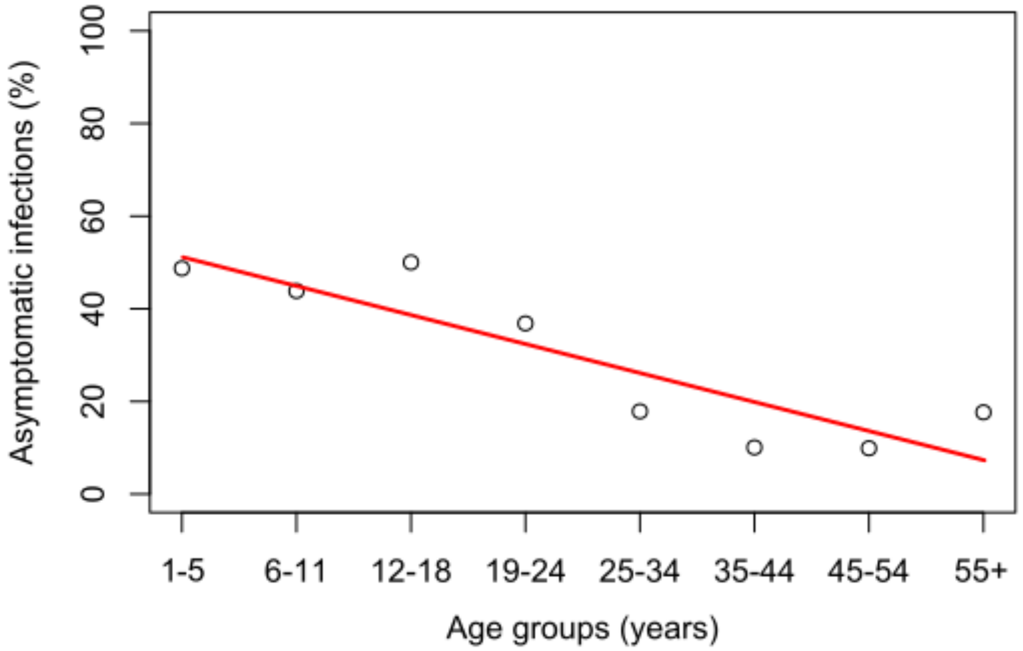

Figure S3 – **Proportion of asymptomatic infections decreases with age**. Line graph demonstrating the decrease in the proportion of asymptomatic infections with increasing age across age-group. Red line indicates line of best fit. Only samples at T1 were included in this analysis (n=1265).

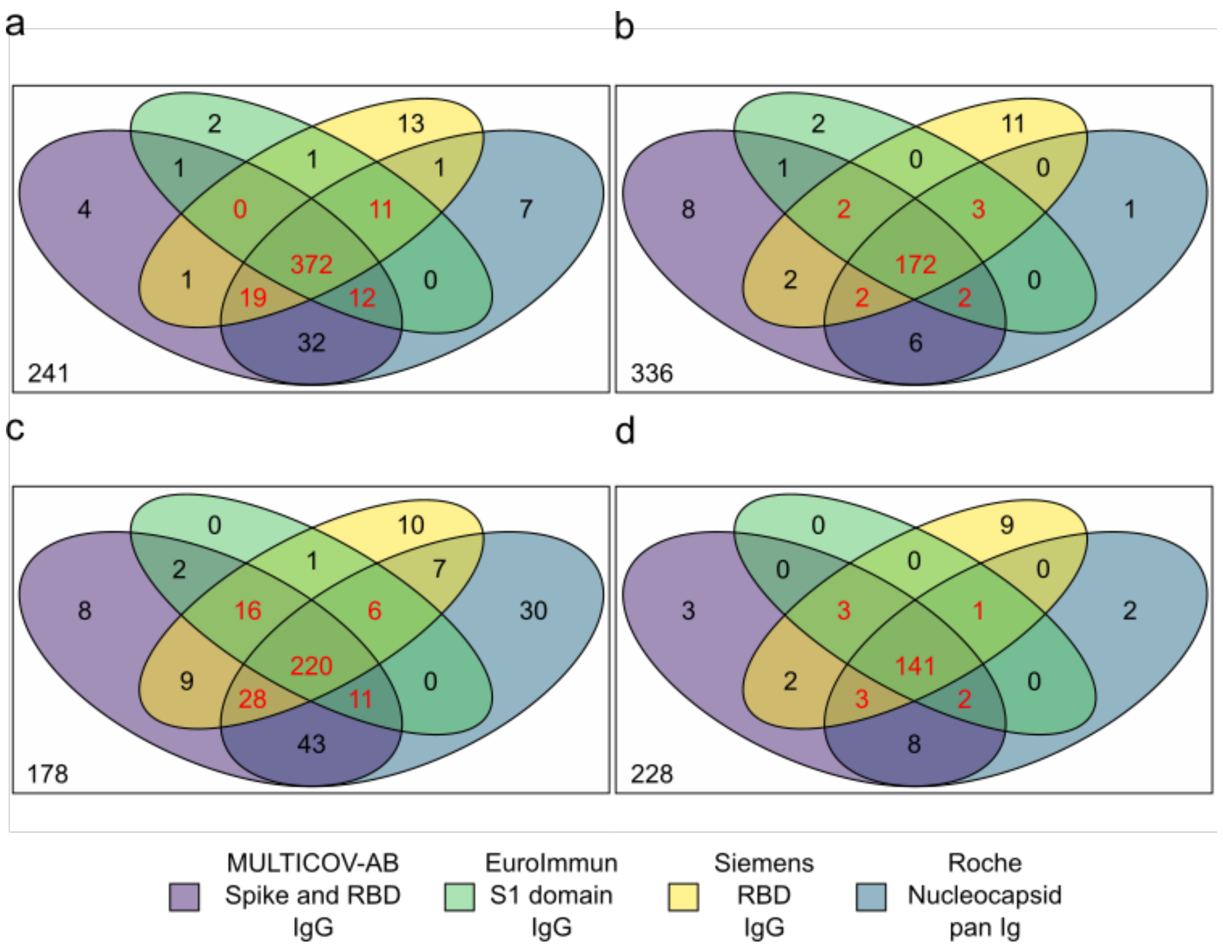

Figure S4 – **Comparative performance of the different serology assays used in this study**. 4-way Venn diagrams showing how each assay classified samples as being seropositive for T1 (a and b) and T2 (c and d) for adults (a and c) and children (b and d). Samples were classified as being positive if three or more assays classified them as being positive (shown in red). Assays are color-coded as defined by the key including the manufacturer or name of the assay, which antigen it uses as a target and which Ig-isotype it measures.

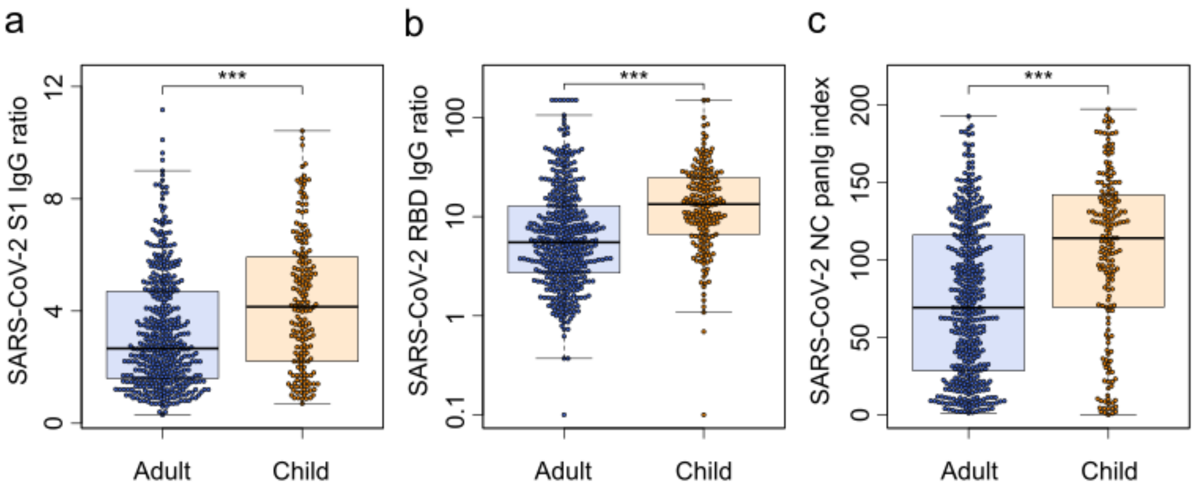

Figure S5 – **Children have higher antibody responses than adults**. Seropositive children (orange, n=181) had significantly higher IgG antibody titres against S1 (a, p<0.001), RBD (b, p<0.001) and nucleocapsid (NC) (c, p<0.001) than seropositive adults (blue, n=414) as determined using the commercial EuroImmun (a), Siemens (b) and Roche (c) assays at T1. Seropositive adults and children were identified using the multi-assay definition of seropositivity explained in the Method section. Box and whisker plots with the box representing the median, 25th and 75th percentiles, while whiskers show the largest and smallest non-outlier values. Outliers were identified using upper/lower quartile ± 1.5 times IQR. Statistical significance was calculated by Mann-Whitney-U (two-sided) with *** indicating a p-value <0.001.

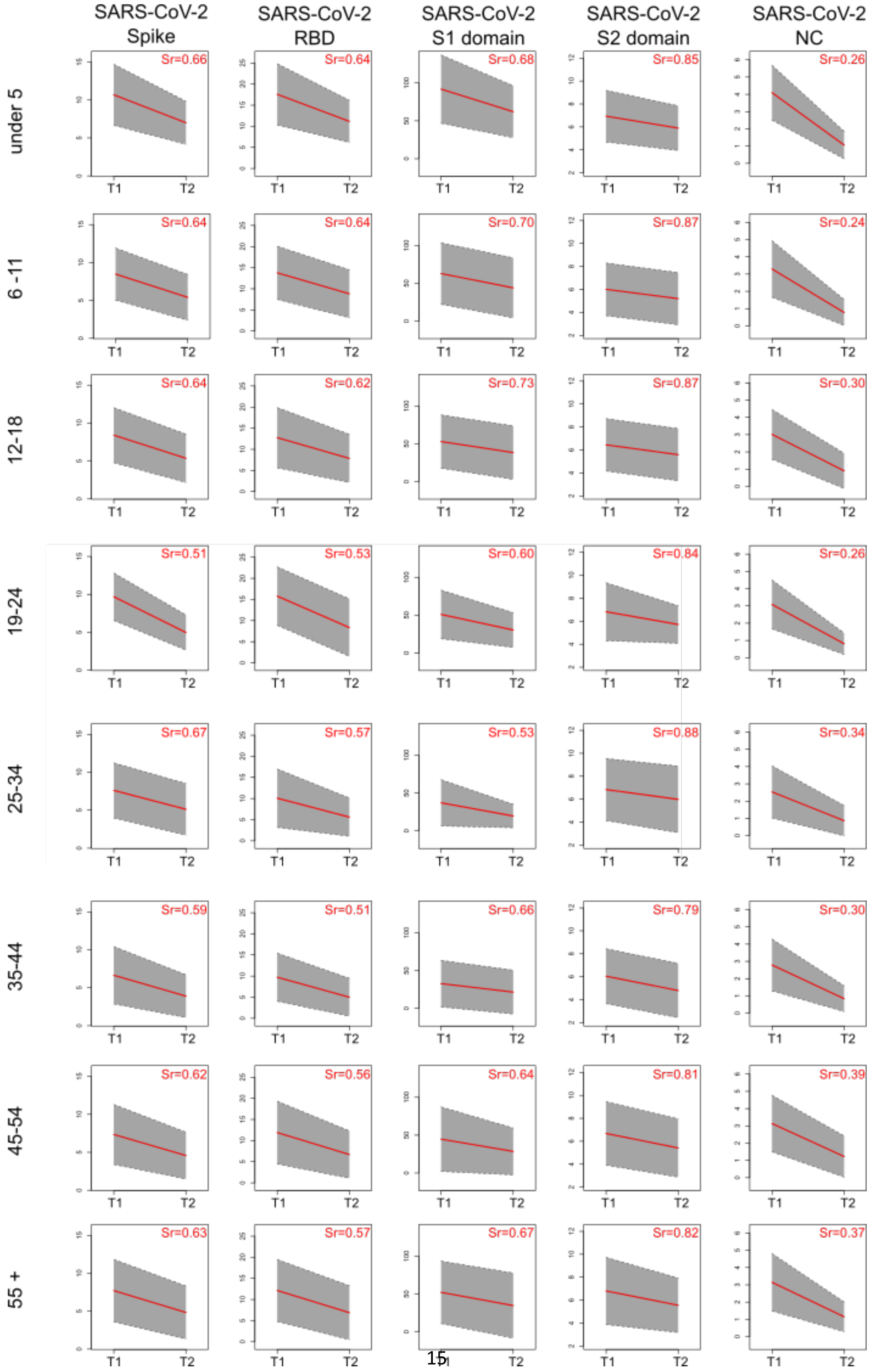

SARS-CoV-2 Spike
SARS-CoV-2 RBD
SARS-CoV-2 S1 domain
SARS-CoV-2 S2 domain
SARS-CoV-2 NC
under 5
Sr=0.66
Sr=0.64
Sr=0.68
Sr=0.85
Sr=0.26
6 - 11
Sr=0.64
Sr=0.64
Sr=0.70
Sr=0.87
Sr=0.24
12-18
Sr=0.64
Sr=0.62
Sr=0.73
Sr=0.87
Sr=0.30
19-24
Sr=0.51
Sr=0.53
Sr=0.60
Sr=0.84
Sr=0.26
25-34
Sr=0.67
Sr=0.57
Sr=0.53
Sr=0.88
Sr=0.34
35-44
Sr=0.59
Sr=0.51
Sr=0.66
Sr=0.79
Sr=0.30
45-54
Sr=0.62
Sr=0.56
Sr=0.64
Sr=0.81
Sr=0.39
55 +
Sr=0.63
Sr=0.57
Sr=0.67
Sr=0.82
Sr=0.37
T1
T2

Figure S6 – **Seroreversion occurs at the same rate in adults and children.** Longitudinal comparison of T1 and T2 samples using MULTICOV-AB to determine the rate of seroreversion. The IgG antibodies against spike trimer, RBD, S1 domain, S2 domain and nucleocapsid (NC) of SARS-CoV-2 are shown. Samples are separated into distinct age groups: under 5 years old (n=28), 6-11 (n=61), 12-18 (n=68), 19-24 (n=14), 25-34 (n=21), 35-44 (n=117), 45-54 (n=148) and over 55 years old (n=31). All y-axis show the normalized MFI. Red lines indicate mean rates of decrease; grey boxes indicate ±1 standard deviation. Sr indicates proportion of signal remaining, calculated as the ratio of the mean MFI at T2 compared to the mean MFI at T1.

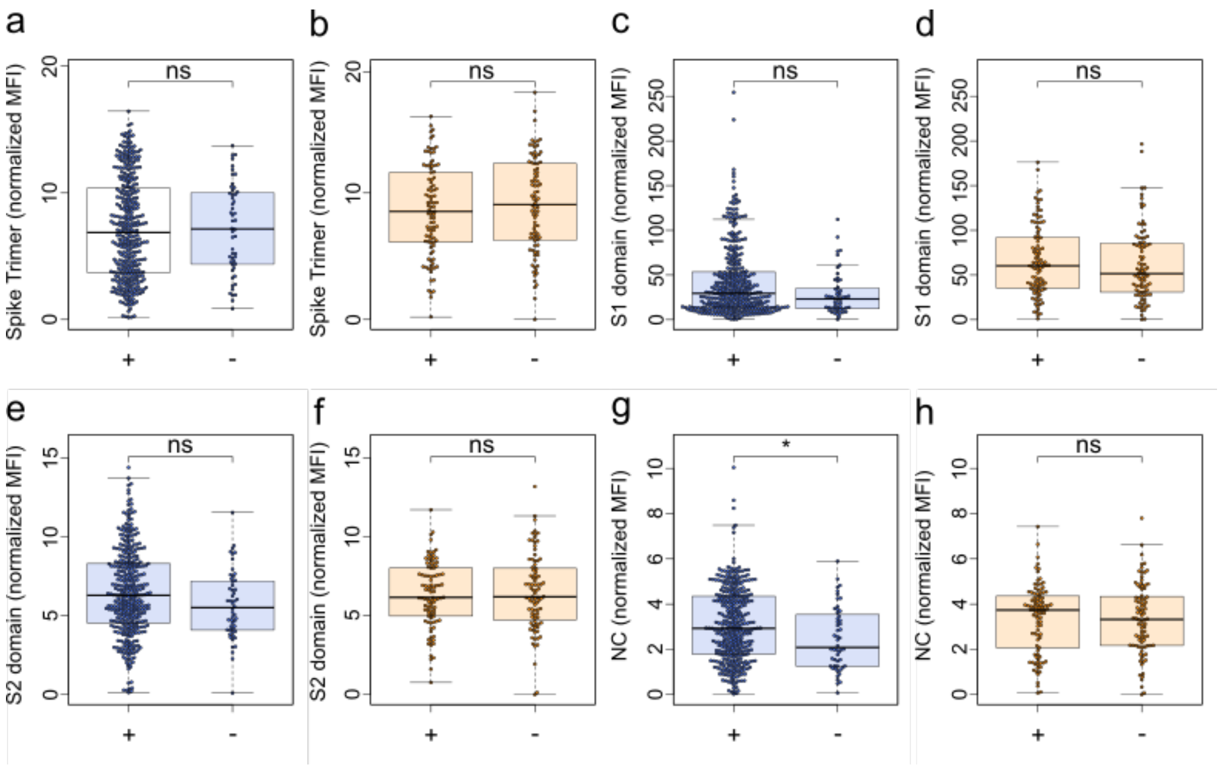

Figure S7 – **There is no difference in antibody response between asymptomatic and symptomatic infections in children**. Box and whisker plots with the box representing the median, 25th and 75th percentiles, while whiskers show the largest and smallest non-outlier values. Outliers were identified using upper/lower quartile ±1.5 times IQR. Statistical significance was calculated by Mann-Whitney-U (two-sided) with * indicating a p-value <0.01. and ns indicating a non-significant p-value >0.01. "+" indicates a symptomatic infection while "-" indicates an asymptomatic infection. There were no significant differences between symptomatic and asymptomatic seropositive children (orange, n=185) in terms of antibody response for the spike trimer (b, p=0.43), S1 domain (d, p=0.34), S2 domain (f, p=0.87) or nucleocapsid (h, p=0.78). Symptomatic and asymptomatic seropositive adults (blue, n=415) showed no significant difference for the spike trimer (a, p=0.94), S1 domain (c, p=0.03) and S2 domain (e, p=0.05), though there was a small significant difference for the nucleocapsid (g, p=0.01).

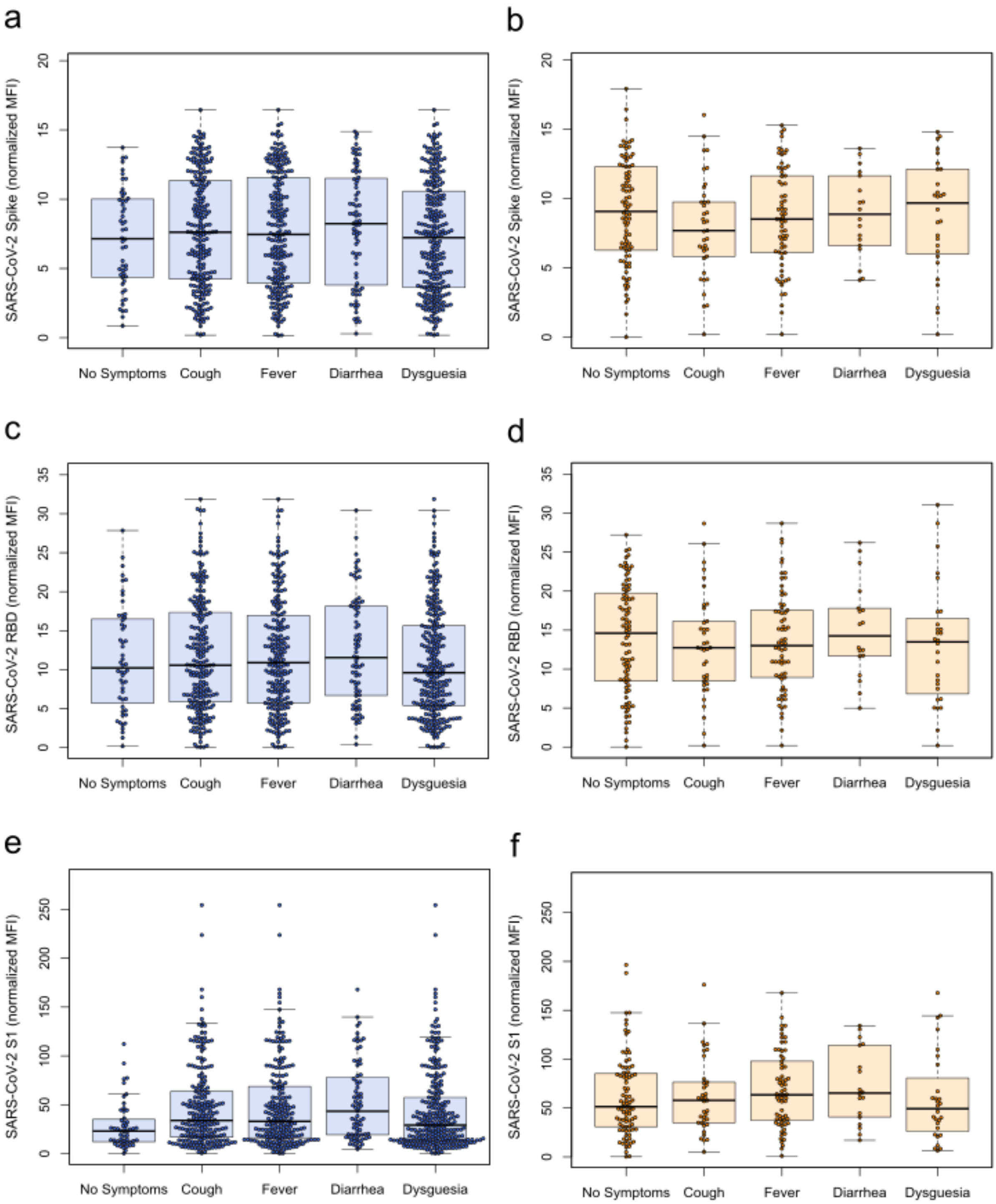

Figure S8 – **Symptoms do not moderate antibody response amongst seropositive individuals after mild COVID-19**. Box and whisker plots with the box representing the median, 25th and 75th percentiles, while whiskers show the largest and smallest non-outlier values. Outliers were identified using upper/lower quartile ±1.5 times IQR. Within the seropositive subgroups, neither adults (a, c, e) nor children (b, d, f) showed any difference in response based on presence of symptoms for either the spike trimer (a and b), RBD (c and d) or S1 domain (e and f) of SARS-CoV-2. Symptom group sizes: no symptoms – adults n=36, children n=83, cough – adults n=221, children n=37, fever – adults n=217, children n=66, diarrhea – adults n=75, children n=18, dysgeusia – adults n=266, children n=28.

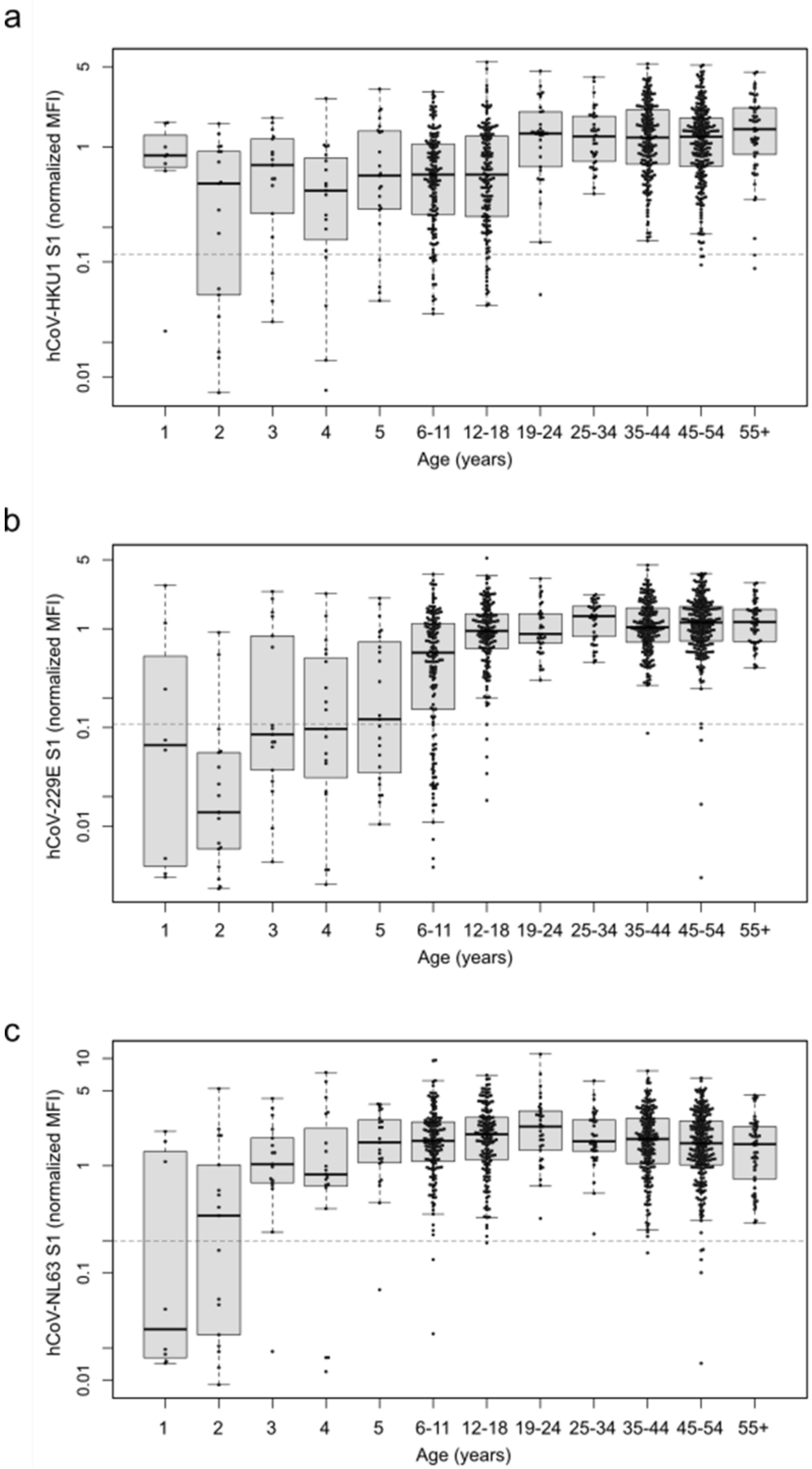
a
hCoV-HKU1 S1 (normalized MFI)
5
1
0.1
0.01
1 2 3 4 5 6-11 12-18 19-24 25-34 35-44 45-54 55+
Age (years)
b
hCoV-229E S1 (normalized MFI)
5
1
0.1
0.01
1 2 3 4 5 6-11 12-18 19-24 25-34 35-44 45-54 55+
Age (years)
c
hCoV-NL63 S1 (normalized MFI)
10
5
1
0.1
0.01
1 2 3 4 5 6-11 12-18 19-24 25-34 35-44 45-54 55+
Age (years)

Figure S9 – **Initial HCoV infection often occurs during the first five years of life**. Box and whisker plots with the box representing the median, 25th and 75th percentiles, while whiskers show the largest and smallest non-outlier values. Outliers were identified using upper/lower quartile ±1.5 times IQR. Single ages from 1 to 5 are shown (1 – n=8, 2 – n=17, 3 – n=17, 4 – n=19, 5 – n=23) with age then grouped into: 6-11 (n=160), 12-18 (n=163), 19-24 (n=34), 25-34 (n=37), 35-44 (n=195), 45-54 (n=346) and over 55 years olds (n=52). For all HCoVs (a – HKU1, b – 229E, c – NL63), the majority of naïve samples are children. Dashed line indicates one-tenth of the mean response of all samples. All samples below the dashed line are considered to be naïve.

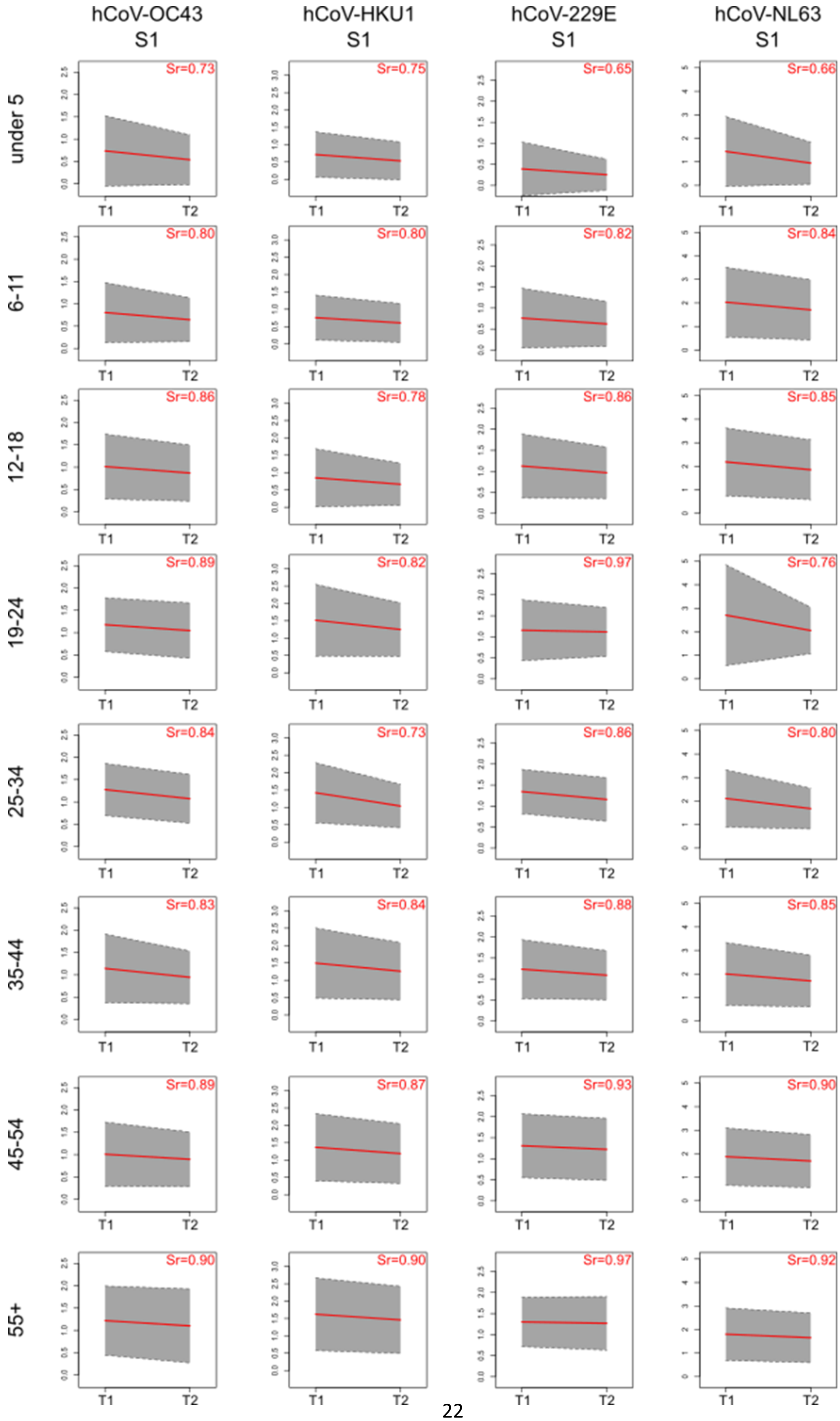
hCoV-OC43
S1
hCoV-HKU1
S1
hCoV-229E
S1
hCoV-NL63
S1
under 5
Sr=0.73
Sr=0.75
Sr=0.65
Sr=0.66
6-11
Sr=0.80
Sr=0.80
Sr=0.82
Sr=0.84
12-18
Sr=0.86
Sr=0.78
Sr=0.86
Sr=0.85
19-24
Sr=0.89
Sr=0.82
Sr=0.97
Sr=0.76
25-34
Sr=0.84
Sr=0.73
Sr=0.86
Sr=0.80
35-44
Sr=0.83
Sr=0.84
Sr=0.88
Sr=0.85
45-54
Sr=0.89
Sr=0.87
Sr=0.93
Sr=0.90
55+
Sr=0.90
Sr=0.90
Sr=0.97
Sr=0.92
T1
T2

Figure S10 – **Children serorevert faster for HCoVs than adults**. Longitudinal comparison of T1 and T2 samples using MULTICOV-AB to determine the rate of seroreversion. The S1 domain of HCoV-OC43, HCoV-HKU1, HCoV-229E and HCoV-NL63 are shown. Samples are separated into distinct age groups: five years old and under (n=84), 6-11 (n=160), 12-18 (n=162), 19-24 (n=32), 25-34 (n=36), 35-44 (n=182), 45-54 (n=228) and over 55 years old (n=48). Red lines indicate mean rate of decrease; grey boxes indicate ±1 standard deviation. Sr indicates the proportion of signal remaining, calculated as the ratio of the mean MFI at T2 compared to the mean MFI at T1.

a

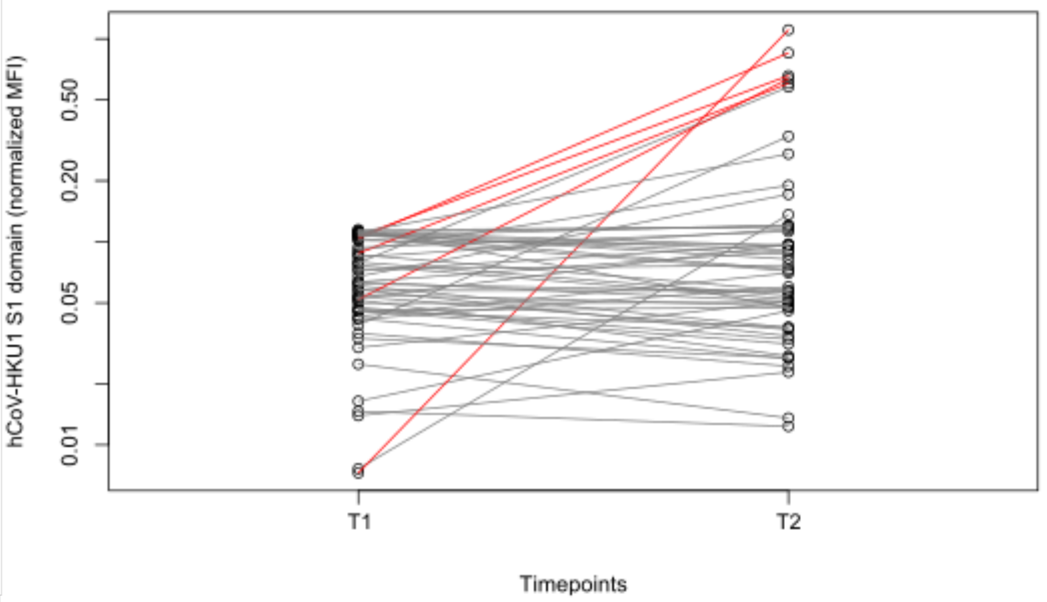
hCoV-HKU1 S1 domain (normalized MFI)
0.01
0.05
0.20
0.50
T1
T2
Timepoints

b

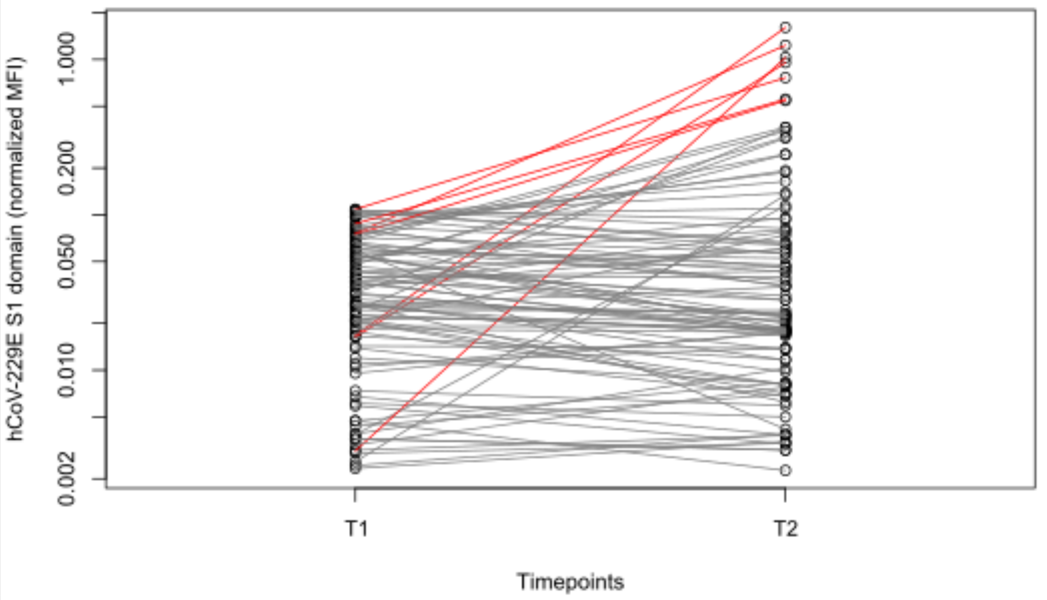
hCoV-229E S1 domain (normalized MFI)
0.002
0.010
0.050
0.200
1.000
T1
T2
Timepoints

c

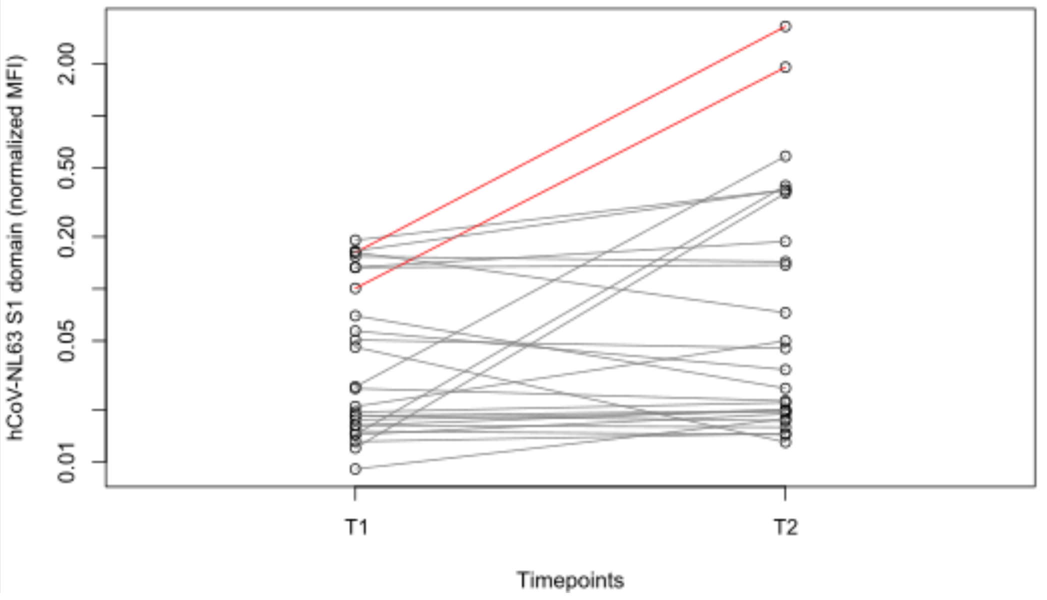
hCoV-NL63 S1 domain (normalized MFI)
0.01
0.05
0.20
0.50
2.00
T1
T2
Timepoints

Figure S11 – **Naïve samples are present within the study, although endemic coronavirus infections persisted during the SARS-CoV-2 pandemic**. Line graphs showing longitudinal antibody response from T1 to T2 for samples defined as naïve at T1. Individuals who remain naïve are shown in grey, individuals who seroconvert between T1 and by T2 are shown in red. Not all individuals who show increased HCoV antibody levels at T2 compared to T1 are considered to have been infected, as some remain within the negative range for the assay at T2. Normalized MFI is shown on a logscale for clarity. Although there is variation in the number of naïve samples between the different HCoVs (a – HKU1, b – 229E, c – NL63), new HCoV infections are seen across all HCoVs.

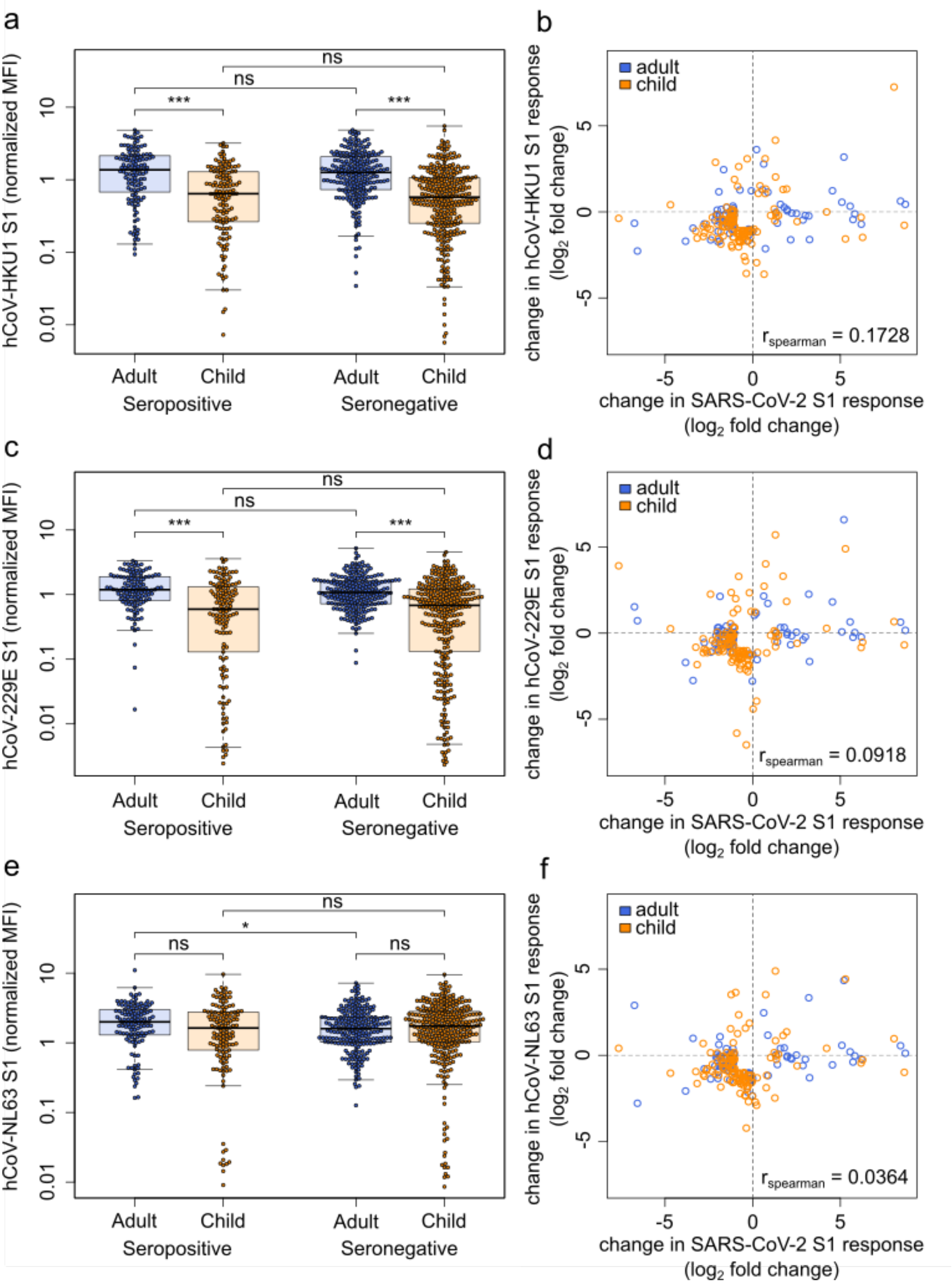

Figure S12 – **HCoVs offer no cross protection towards SARS-CoV-2, nor do they show a boost-back antibody response following SARS-CoV-2 infection**. Samples from households with a known index case were examined with MULTICOV-AB to determine whether the antibody response to endemic coronaviruses (HCoV) provides any protection against SARS-CoV-2. (a, c and e) Box and whisker plots demonstrating no significant difference between SARS-CoV-2 seropositive and seronegative adults (n=440) or children (n=436) in terms of HCoV-HKU1 (a, adult p=0.67, child p=0.47) or HCoV-229E (c, adult p=0.14, child p=0.99). For HCoV-NL63, there was a small significant difference for adults only (e, adults p=0.009, children p=0.35). Boxes represent the median, 25th and 75th percentiles, and whiskers show the largest and smallest non-outlier values. Outliers were identified using upper/lower quartile ± 1.5 times IQR. Statistical significance was calculated by Mann-Whitney-U (two-sided) with *** indicating a p-value <0.001, * indicating a p-value <0.01 and ns indicating a p-value >0.01 (b, d and f). When comparing paired samples longitudinally within the SARS-CoV-2 seropositive subgroup, there was no association between change in SARS-CoV-2 antibody level and change in HCoV antibody level for HCoV-HKU1 (b), HCoV-229E (d) or HCoV-NL63 (f)  and in either adults (b - n=79, d – n=74, f – n=80) or children (b -n=117, d – n=114, f – n=118). Change in response is presented as log2-fold change from T1 to T2 and only samples with a log2-fold change > 1 or < -1 are shown. Spearman’s rank was used to calculate correlations between the changes in HCoV antibody level and change in SARS-CoV-2 antibody level.

#### Supplementary Tables

Table S1 – **Comparative seroprevalence between different assays used in the study**

| Assay | Seropositive | Seronegative | Total | % of Seropositive |
|---|---|---|---|---|
| EuroImmun S1 IgG | 991 | 1245 | 2236 | 44.32 |
| Roche Elecsys N pan Ig | 1149 | 1087 | 2236 | 51.39 |
| Siemens RBD IgG | 1067 | 1169 | 2236 | 47.72 |
| MULTICOV S and RBD IgG | 1142 | 1094 | 2236 | 51.07 |

Only samples that were measured with all four assays are considered. For each assay, the manufacturer or name of the assay is stated, as well as the target antigen and Ig-isotype detected.

Table S2 – **Symptom frequency and diagnostic performance in children under 18**

| | | Age group | | | | | | All children | |
|---|---|---|---|---|---|---|---|---|---|
| | | Under 5 | | 6 to 11 | | 12 to 18 | | | |
| *Symptom* | Present or Absent | n (%) | Seropositive (PPV, 95% CI) | n (%) | Seropositive (PPV, 95% CI) | n (%) | Seropositive (PPV, 95% CI) | n (%) | Seropositive (PPV, 95% CI) |
| Fever | Present | 37 (28.03) | 17 (0.46, 0.33-0.59) | 46 (21.70) | 28 (0.61, 0.48-0.72) | 28 (13.40) | 21 (0.75, 0.57-0.87) | 111 (20.07) | 66 (0.59, 0.51-0.67) |
| | Absent | 95 (71.97) | 23 (0.24, 0.19-0.30) | 166 (78.30) | 45 (0.27, 0.24-0.31) | 181 (86.60) | 53 (0.29, 0.27-0.32) | 442 (79.93) | 121 (0.27, 0.25-0.29) |
| Cough | Present | 28 (21.21) | 9 (0.32, 0.19-0.48) | 43 (20.28) | 14 (0.33, 0.21-0.46) | 28 (13.40) | 14 (0.50, 0.34-0.66) | 99 (17.90) | 37 (0.37, 0.29-0.46) |
| | Absent | 104 (78.79) | 31 (0.30, 0.25-0.35) | 169 (79.72) | 59 (0.35, 0.31-0.38) | 181 (86.60) | 60 (0.33, 0.30-0.36) | 454 (82.10) | 150 (0.33, 0.31-0.35) |
| Diarrhea | Present | 8 (6.06) | 3 (0.38, 0.13-0.71) | 17 (8.02) | 11 (0.65, 0.41-0.83) | 8 (3.83) | 4 (0.50, 0.20-0.80) | 33 (5.97) | 18 (0.55, 0.38-0.70) |
| | Absent | 124 (93.94) | 37 (0.30, 0.27-0.32) | 195 (91.98) | 62 (0.32, 0.30-0.34) | 201 (96.17) | 70 (0.35, 0.33-0.36) | 520 (94.03) | 169 (0.33, 0.31-0.34) |
| Dysguesia | Present | 1 (0.76) | 1 (1.00, n/a) | 7 (3.30) | 6 (0.86, 0.42-0.98) | 24 (11.48) | 21 (0.88, 0.68-0.96) | 32 (5.79) | 28 (0.88, 0.71-0.95) |
| | Absent | 131 (99.24) | 39 (0.30, 0.29-0.30) | 205 (69.70) | 67 (0.33, 0.32-0.34) | 185 (88.52) | 53 (0.29, 0.27-0.31) | 521 (94.21) | 159 (0.31, 0.30-0.31) |

The frequency of each symptom within the study population, shown number of individuals (n, also as %) either with (present) or without (absent) this symptom, and the number of individuals (n, also as  %) within these groups who were seropositive for SARS-CoV-2. Children are split into three groups: under 5-year olds (n=132), 6- to 11-year olds (n=212) and 12- to 18-year olds (n=209). Positive Predictive Value (PPV) for seropositivity in the presence or absence of each symptom, 95% Confidence Intervals (CI) are the standard logit confidence intervals calculated as given by Mercaldo et al. 2007[5].

Table S3 – **List of antigens used in MULTICOV-AB**

| Disease | Antigen | Manufacturer | Cat. No. |
|---|---|---|---|
| SARS-CoV-2 | Spike Trimer | NMI | - |
| SARS-CoV-2 | RBD | NMI | - |
| SARS-CoV-2 | S1 domain | NMI | - |
| SARS-CoV-2 | S2 domain | Sino | 40590 |
| SARS-CoV-2 | Nucleocapsid | Aalto | 6404-b |
| SARS-CoV-2 | Nucleocapsid N-terminal domain | NMI | - |
| SARS-CoV-2 | RBD alpha variant | NMI | - |
| SARS-CoV-2 | RBD beta variant | NMI | - |
| hCoV-OC43 | S1 domain | NMI | - |
| hCoV-OC43 | Nucleocapsid | NMI | - |
| hCoV-OC43 | Nucleocapsid N-terminal domain | NMI | - |
| hCoV-HKU1 | S1 domain | NMI | - |
| hCoV-HKU1 | Nucleocapsid | NMI | - |
| hCoV-HKU1 | Nucleocapsid N-terminal domain | NMI | - |
| hCoV-NL63 | S1 domain | NMI | - |
| hCoV-NL63 | Nucleocapsid | NMI | - |
| hCoV-NL63 | Nucleocapsid N-terminal domain | NMI | - |
| hCoV-229E | S1 domain | NMI | - |
| hCoV-229E | Nucleocapsid | NMI | - |
| hCoV-229E | Nucleocapsid N-terminal domain | NMI | - |
| CONTROL | Human IgG | NMI | - |
| CONTROL | Goat-anti-human IgG | NMI | - |
| CONTROL | Human IgA | NMI | - |
| CONTROL | Goat-anti-human IgA | NMI | - |

List of antigens included in MULTICOV-AB in this study, including information about their manufacturer, and if available, their category number. Full information on the NMI produced antigens were used as previously published[1,2].
